# Germline and Somatic Mutations in DNA Methyltransferase 3A *(DNMT3A)* Predispose to Pulmonary Arterial Hypertension (PAH) in Humans and Mice: *Implications for Associated PAH*

**DOI:** 10.1101/2023.12.30.23300391

**Authors:** Ruaa Al-Qazazi, Isaac M. Emon, François Potus, Ashley Y. Martin, Patricia D.A. Lima, Caitlyn Vlasschaert, Kuang-Hueih Chen, Danchen Wu, Asish Das Gupta, Curtis Noordhof, Lindsay Jefferson, Amy J. M. McNaughton, Alexander G. Bick, Michael W. Pauciulo, William C. Nichols, Wendy K. Chung, Paul M. Hassoun, Rachel L. Damico, Michael J. Rauh, Stephen L. Archer

**Affiliations:** Department of Medicine, Queen’s University, Kingston, Ontario, Canada; Pulmonary Hypertension Research Group, Institut Universitaire de Cardiologie et de Pneumologie de Québec Research Center, Laval University, Quebec City, Canada; Queen’s Cardiopulmonary Unit, Queen’s University, Kingston, Ontario, Canada; Department of Pathology and Molecular Medicine, Queen’s University, Kingston, Ontario, Canada; Department of Medicine, Vanderbilt University Medical Center, Nashville, Tennessee, USA; Division of Human Genetics, Cincinnati Children’s Hospital Medical Center, and Department of Pediatrics; University of Cincinnati College of Medicine, Cincinnati, Ohio, USA; Department of Pediatrics, Boston Children’s Hospital, Harvard Medical School, Boston, Massachusetts, USA; Department of Medicine, Division of Pulmonary and Critical Care Medicine, Johns Hopkins University, Baltimore, Maryland, USA

**Keywords:** clonal hematopoiesis of indeterminate potential (CHIP), hypoxia, associated pulmonary arterial hypertension (APAH), epigenetics, inflammation, DNA methylation, scleroderma

## Abstract

**Background:** Mutations are found in 10-20% of idiopathic PAH (IPAH) patients, but none are consistently identified in connective tissue disease-associated PAH (APAH), which accounts for ∼45% of PAH cases. *TET2* mutations, a cause of clonal hematopoiesis of indeterminant potential (CHIP), predispose to an inflammatory type of PAH. We now examine mutations in another CHIP gene, *DNMT3A*, in PAH.

**Methods:** We assessed *DNMT3A* mutation prevalence in PAH Biobank subjects as compared with controls, first using whole exome sequencing (WES)-derived CHIP calls in 1832 PAH Biobank patients versus 7509 age-and sex-matched gnomAD controls. We then performed deep, targeted panel sequencing of CHIP genes on a subset of 710 PAH Biobank patients and compared the prevalence of *DNMT3A* mutations therein to an independent pooled control cohort (N = 3645). In another cohort of 80 PAH patients and 41 controls, *DNMT3A* mRNA expression was studied in peripheral blood mononuclear cells (PBMCs). Finally, we evaluated the development of PAH in a conditional, hematopoietic, *Dnmt3a* knockout mouse model.

**Results:** *DNMT3A* mutations were more frequent in PAH cases versus control subjects in the WES dataset (OR 2.60, 95% CI: 1.71-4.27). Among PAH patients, 33 had *DNMT3A* variants, most of whom had APAH (21/33). While 21/33 had somatic mutations (female:male 17:4), germline variants occurred in 12/33 (female:male 11:1). Hemodynamics were comparable with and without *DNMT3A* mutations (mPAP=58±21 vs. 52±18 mmHg); however, patients with *DNMT3A* mutations were unresponsive to acute vasodilator testing. Targeted panel sequencing identified that 14.6% of PAH patients had CHIP mutations (104/710), with *DNMT3A* accounting for 49/104. There was a significant association between all CHIP mutations and PAH in analyses adjusted for age and sex (OR 1.40, 95% CI: 1.09-1.80), though *DNMT3A* CHIP alone was not significantly enriched (OR:1.15, 0.82-1.61). *DNMT3A* expression was reduced in patient-derived versus control PAH-PBMCs. Spontaneous PAH developed in *Dnmt3a^-/-^* mice, and it was exacerbated by 3 weeks of hypoxia. *Dnmt3a^-/-^* mice had increased lung macrophages and elevated plasma IL-13. The IL-1β antibody canakinumab attenuated PAH in *Dnmt3a^-/-^* mice.

**Conclusions:** Germline and acquired *DNMT3A* variants predispose to PAH in humans. *DNMT3A* mRNA expression is reduced in human PAH PBMCs. Hematopoietic depletion of Dnmt3a causes inflammatory PAH in mice. *DNMT3A* is a novel APAH gene and may be a biomarker and therapeutic target.

## Introduction

Pulmonary arterial hypertension (PAH) is a pulmonary vasculopathy characterized by an elevation in mean pulmonary arterial pressure (mPAP) >20 mmHg, pulmonary vascular resistance (PVR) >3 Wood Units and normal pulmonary capillary wedge pressure (PCWP) ≤15 mmHg (in the absence of significant left heart disease, lung disease, or thromboembolic disease)^1^. There are different subtypes of PAH, including associated PAH (APAH), heritable PAH (HPAH), comprising familial and sporadic cases, and idiopathic PAH (IPAH), where no identifiable cause is identified. While gene mutations have been identified in 70% of patients with familial PAH and 10-20% of IPAH cases, causal mutations are rarely identified in APAH^2^. According to the Pulmonary Hypertension Association Registry (PHAR), the prevalence of PAH by cause is: idiopathic (41%), heritable (3%), connective tissue disease (32%), drug-induced (11%), portopulmonary hypertension (7%), congenital heart disease (5%), HIV (2%), and pulmonary veno-occlusive disease (1%). The percentage of adult patients with PAH that have APAH exceeds 45%^3^. APAH mostly occurs in patients with underlying connective tissue diseases, such as scleroderma, the preponderance of whom are women^4^. However, no specific gene mutations have been consistently associated with APAH. The etiology of APAH is multifactorial, involving complex interactions between genetic, environmental, and immunological factors, leading to excessive inflammation. The search for PAH risk genes is important since the underlying mechanisms driving PAH pathogenesis in many patients remain unclear^5^. Most mutations in PAH are germline mutations that are inherited by an autosomal dominant mechanism with variable penetrance^6^. The first identified PAH gene, *BMPR2* remains the most prevalent of the ∼20 PAH related genes^7,8^. Identifying PAH-predisposing genes informs personal risk assessment, assists people in their family planning, and enables early PAH detection through targeted screening of high-risk individuals. The known PAH genes do not explain the inflammatory form of PAH found in people with APAH.

The primary objective of this research is to examine the impact of mutations in the gene encoding DNA (cytosine-5) methyltransferase 3 alpha (*DNMT3A*), a major epigenetic regulator that mediates *de novo* DNA methylation, on the risk of developing PAH. To achieve this, four studies were performed. First, gene-specific, rare variant association analyses were conducted using a dataset comprising 1832 patients in the PAH Biobank who had undergone whole exome sequencing (WES)^9^. Second, targeted panel sequencing was subsequently performed on 710 PAH Biobank patients to obtain a deeper, more sensitive identification of *DNMT3A* mutations. Third, in an independent cohort of 80 patients and 41 controls, we measured the expression levels of *DNMT3A* mRNA in peripheral blood mononuclear cells (PBMCs). Fourth, we utilized a mouse model in which we conditionally depleted *Dnmt3a* expression specifically in hematopoietic lineage cells (*Dnmt3a*^-/-^ mice) and examined the development of PAH at 4.5 and 9 months.

Somatic *DNMT3A* mutations in hematopoietic stem cells promote clonal hematopoiesis of indeterminate potential (CHIP), a precursor to hematological malignancies, such as myeloproliferative and myelodysplastic neoplasms, and acute myeloid leukemia^10,11,12^. Mutations in genes encoding DNA methylation enzymes, including DNA methyltransferases (*DNMTs*), and demethylation mediator enzymes, such as Tet methylcytosine dioxygenase 2 (*TET2*), result in aberrant DNA hypo-or hyper-methylation, respectively, leading to inflammation^13–14^. The loss of function of DNMT3A and TET2 was also found to trigger a type 1 interferon response in macrophages due to impaired mitochondrial DNA integrity^15^. CHIP mutations promote inflammation in vascular diseases, like atherosclerosis, hemorrhagic and ischemic stroke^16,17,1819^. *TET2* mutations promote PAH with inflammation that is more severe than is seen in PAH patients lacking *TET2* mutations^20^. PAH has many similarities to cancer and inflammatory diseases, making *DNMT3A* an attractive candidate PAH gene^21^.

This study identified an increased incidence of deleterious germline and somatic variants in *DNMT3A*, predominantly in females with APAH. Targeted panel sequencing of common CHIP genes shows that CHIP gene mutations in general are more common among individuals with PAH, with *DNMT3A* being the most often mutated CHIP gene. Moreover, expression of *DNMT3A* mRNA was reduced in PBMCs from PAH versus control subjects. Finally, *Dnmt3a*^-/-^ mice developed spontaneous PAH that was characterized by lung infiltration with inflammatory cells, specifically macrophages. The discovery of *DNMT3A* variants that are predicted to disrupt gene function and lower expression of DNMT3A in the blood of PAH patients adds to our understanding of the complex genetic landscape of PAH and highlights the significance of *DNMT3A* as a precipitant of APAH. The PAH susceptibility of mice lacking *Dnmt3a* in their hematopoietic system confirms the biologic relevance of this pathway to PAH. This research identifies *DNMT3A* as a PAH-inducing CHIP gene and provides valuable insights into the role of inflammation in the pathogenesis of PAH. DNMT3A may be a useful biomarker and target for therapeutic interventions in PAH.

## Methods

The authors declare that all supporting data are available within the article and the online Supplemental Data.

### 1- Human studies

#### 1.1 Study subjects

Patient IDs were not known to anyone outside the research group. The genetic sequencing data cohort is comprised of subjects from The National Biological Sample and Data Repository for PAH (aka PAH Biobank). The PAH Biobank is maintained at Cincinnati Children’s Hospital Medical Center (CCHMC) and is comprised of biological samples, clinical data, and genetic data obtained from over 2800 PAH patients enrolled from 38 North American PH Centers. Ethical approval of this study was given by the Cincinnati Children’s Hospital Medical Center (lead centre) Institutional Review Board, while original Biobank data collection was approved by individual Enrolling Centers IRB. The samples and data are made available for use by the PAH research community, including this study, as approved by the lead centre, Cincinnati Children’s Hospital Medical Center IRB.

#### 1.2 Exome sequencing and genetic case-control comparisons

We compared the rate of pathogenic *DNMT3A* variants and average age of the cases in the PAH Biobank to the SPARK and gnomAD databases (total cases in the databases were: 2707, 11,198, 141,456 respectively; **Figure 1**). Patients in the PAH Biobank were excluded if they had a known PAH gene mutation. The pathogenic variant rate was compared between groups using Fisher’s Exact Test. A p<0.05, after correction for multiple comparisons, was considered statistically significant. DNA was extracted from whole blood of participants in the National Biological Sample and Data Repository for PAH (PAH Biobank). Whole Exome Sequencing (WES) was carried out in collaboration with the Regeneron Genetics Center and/or at the Cincinnati Children’s Hospital Medical Center DNA Sequencing and Genotyping Core, as described^22,23^. We used: 1) Burrows-Wheeler Aligner (BWA-MEM) to map and align paired-end reads to the human reference genome (GRCh38)^24^, 2) Picard MarkDuplicates to identify and flag PCR duplicates, and 3) GATK HaplotypeCaller to call genetic variants^25^. We used ANNOVAR^26^ to aggregate variant annotations, including population allele frequency (gnomAD^27^ exome/genome, ExAC, and 1000 genomes). We predicted functional effects based on RefSeq and predicted deleterious scores of missense variants using REVEL.

**Figure 1:**
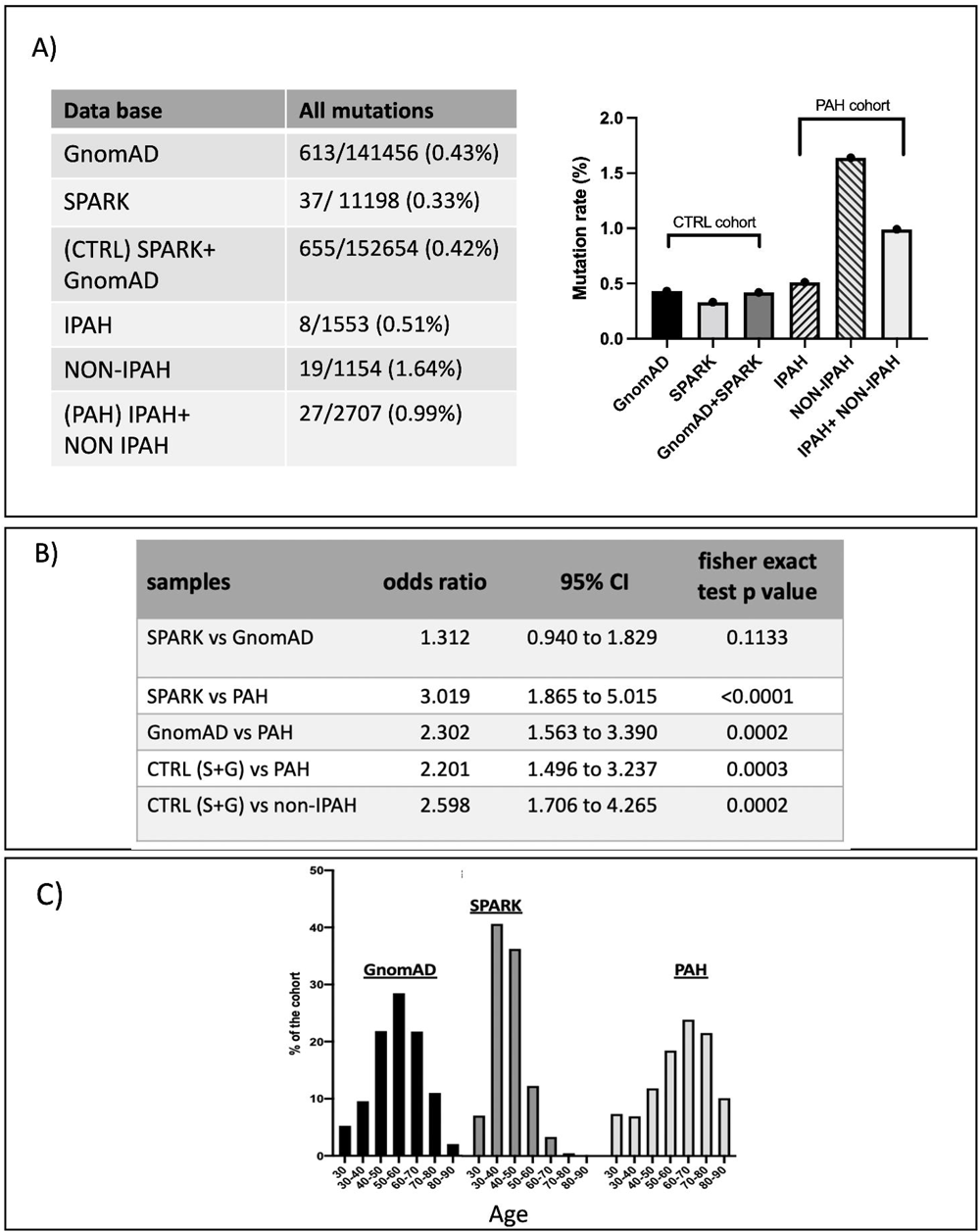
Prevalence of DNMT3A Mutations in PAH Patients using Whole Exome Sequencing: A Comparative Analysis with GnomAD and SPARK Databases. **A)** *DNMT3A* mutation rates are shown for the gnomAD and SPARK control databases individually, and for the SPARK and gnomAD databases combined. Mutation rates are also shown for PAH, IPAH and NON-IPAH patients. Mutation rate is shown on the y-axis of the graph (in %) and is displayed for the 3 control cohorts and 3 PAH cohorts**. B)** Fisher’s exact test was used to compare the mutation rate between PAH, SPARK and gnomAD databases. Fisher’s exact test p-values as well as odds ratios and the associated confidence intervals (95% confidence) are shown for several PAH vs control comparisons. Mutation rates were significantly increased in the PAH Biobank (1%) compared to both SPARK (0.33%; p=<0.0001) and gnomAD (=> 0.43%; p*=*0.0002) controls. Mutation rates were also significantly increased in both total PAH patients and non-IPAH patients (1.64%) compared to both control databases combined (0.42%, p=0.0003 and 0.0002, respectively). There was no significant difference between the SPARK and gnomAD control databases (p=0.1133). **C)** The age groups of the cohorts were more comparable between gnomAD and PAH databases. % of the cohort (y-axis) at each age range (x-axis) is shown for the PAH Biobank as well as both control databases.

1832 unrelated European PAH cases were identified in the PAH Biobank. Rare variants were defined as variants with an allele frequency <0.01% across all gnomAD exome sequencing samples (European and non-European). Heuristic filters were used to minimize technical artifacts between cases and controls and to exclude somatic mutations at lower variant allele fractions (VAFs). Variants that met any of the following criteria were excluded: missingness >10%, minimum alternate allele read depth ≤4 reads, alternative allele fraction ≤25%, or genotype quality <90. We used European gnomAD whole genome sequence (WGS) data for 7509 age and sex-matched individuals as the control set. Only variants with FILTER “PASS” in gnomAD WGS (data release v 2.02) and located in the IDT xGen captured protein coding region were included in the analysis. Finally, we observed that the frequency of rare synonymous variants in cases and controls was virtually identical, indicating that the data sets are comparable (enrichment rate=1.0, p=0.11).

Assuming linkage disequilibrium is negligible among ultra-rare variants, we performed gene-wise burden tests of rare variants for *DNMT3A* using exact binomial tests. We tested the association in two groups of variants between cases and controls: (a) rare deleterious missense variants (“D-MIS”, defined as REVEL score >0.5); (b) rare D-MIS or likely-gene-disrupting (LGD) variants (predicted loss of function-frameshift, stopgain, stoploss, splice). We then assessed the PAH Biobank cohort of 2572 cases who were free from other known PAH-associated gene variants for rare (AF<0.0001), deleterious (LGD or D-MIS) variants in *DNMT3A* using Integrative Genome Viewer (Illumina, San Diego, CA). Nearly 100% of both cases and controls had >10x sequencing coverage across the *DNMT3A* gene, and 90% had >15x coverage across most of the targeted regions (as previously reported)^28^. All *DNMT3A* variants were confirmed by Sanger sequencing of exonic PCR products. To detect likely somatic variants (mosaicism) in *DNMT3A*, we first used SAMtools (version 1.3.1-42)^29^ to improve calling of genetic variants with low allele fraction. We then processed SAMtools calls using a set of heuristic filters to remove variants located in repeat regions (mappability, segmental duplication) or showing evidence of strand bias, and screened all variants using Integrative Genome Viewer. We then took the union of variants called by GATK HaplotypeCaller and SAMtools and considered variants with alternative allele fraction less than 25% as likely somatic mutations. Somatic mutations in the PAH Biobank were confirmed by TA cloning (Thermo Fisher, Waltham, MA) of exonic PCR products followed by Sanger sequencing of individual clones. Details are available in the online supplements.

#### 1.3 Targeted panel sequencing and case-control comparisons

A subset of 710 samples from the PAH biobank were sequenced at Vanderbilt University Medical Center (VUMC) using a targeted gene panel covering the most frequently mutated CHIP genes^30^. CHIP variants were identified using standard methodology for this panel, as described^30^. A pooled control cohort (total N = 3645) was defined, comprised of 244 individuals who were older relatives of individuals with developmental disorders (samples collected for trio exome analysis) and 3401 individuals from the VUMC BioVU biorepository who were initially sequenced as controls for a different study on the basis of not having chronic kidney disease but otherwise not selected for any other clinical characteristics. The control cohort was sequenced on the same platform and CHIP variants were identified using the same method as PAH samples. The prevalence of CHIP in PAH cases compared to controls was evaluated using logistic regression, adjusting for participant age and sex.

#### 1.4 Microarray and Gene Expression

RNA was extracted from peripheral blood mononuclear cells (PBMCs) of 50 patients with scleroderma-associated PAH (SSc-PAH), 30 patients with idiopathic PAH (IPAH), 19 scleroderma patients without PAH (SSc), and 41 healthy controls (140 patients total), at Johns Hopkins University, as previously described^31^. Gene expression microarray assays were conducted on the extracted RNA samples following published protocols^31^. The resulting expression data were deposited in the National Center for Biotechnology Information’s Gene Expression Omnibus (GEO) and can be accessed using the GEO Series accession number GSE33463. The analytical methods are published^31^. All cases had adult-onset PAH, with genetically determined ancestries as follows: 82.1% European, 14.3% African, and smaller percentages of other origins. The sex ratios (female: male) were as follows: control 4.9:1, IPAH 5:1, SSc-PAH 3.7:1. Cohort data previously published in a study focused on *TET2* by *Potus et al*^28^. We examined the expression of DNMT3A transcript variants 2 (NM_153759.3), 3 (NM_022552), and 4 (NM_175630.1), which code for the three protein isoforms DNMT3A2, DNMT3A3, and DNMT3A4, respectively^32^ and performed a receiver operating characteristic (ROC) analysis.

### 2 Animal studies

Rodent experiments were conducted following the Canadian Council on Animal Care (CCAC) regulations approved by Queen’s University Animal Care Committee (Protocol # 2021-2128 and # 2022-2130).

#### 2.1 *DNMT3A* knockout mouse model

We produced male and female conditional hematopoietic *Dnmt3a* knockout mice by crossing parental floxed and Vav-iCre mice as described by the Jackson Laboratory (https://www.jax.org/strain/008610) and by *Joseph et al.*^3334^This knockout simulates the ‘loss of function’ effects of *DNMT3A* somatic mutations seen in patients with PAH in our U.S. PAH Biobank data. To confirm genotype, [controls (*Dnmt3a*^f/f^) vs heterozygous (*Dnmt3a*^+/-^) vs. full knockout (*Dnmt3a^−/−^*)], we performed polymerase chain reaction (PCR) genotyping of PBMCs, as well as ear tissue, to ensure no germline knockout, following manufacturer’s protocol (Accustart II PCR Genotyping Kit”: Cat# 95135-500 – QuantaBio – Beverly MA). PCR product was loaded on a 2% agarose gel stained with SybrSafe and run at 80V for 30 minutes prior to imaging on ChemiDocTMMP imaging system (Bio-Rad Laboratories; Mississauga, ON, Canada). Mice were kept in normoxic conditions (∼21% O_2_) to measure the spontaneous development of PAH over 9 months. To accelerate the rate at which PAH develops, a group of mice (3-month-old) were exposed to a second hit (hypoxia; 10% O_2_; 3 weeks) followed by 3 weeks of normoxia. This group exposed to hypoxia was compared to age and sex matched mice who were handled identically but were not exposed to hypoxia. The end point studies were conducted when both groups were 4.5-months of age. Hypoxia alone can induce group 3 pulmonary hypertension; thus, mice are maintained 3 weeks in normoxia to allow for recovery from any non-Group 1 PH development.

#### 2.2 Non-invasive heart function assessment using cardiac ultrasound

Non-invasive doppler, 2-dimensional (2-D), and time-motion (M-mode) high frequency ultrasound was performed using (Vevo 2100, Visual Sonics, Toronto, ON, Canada) and a 37 MHz probe^35^. Anaesthesia was induced with 4% inhaled isoflurane and maintained with 2% isoflurane for the duration of the procedure. M-mode and 2-D mode were used to measure: pulmonary artery acceleration time (PAAT), main pulmonary artery (PA) inner diameter at the level of the pulmonary outflow tract during mid-systole, tricuspid, and mitral annular plane systolic excursion (TAPSE and MAPSE). Cardiac output (CO) was estimated (HR X VTI X 3.14 X radius^2^), where HR is heart rate, VTI is systolic velocity time integral over the main pulmonary artery flow (obtained from pulsed-wave Doppler), and 3.14 X radius^2^ is the area of the main PA at mid-systole.

#### 2.3 RV and LV pressure-volume loop assessment by closed and open-chest catheterization

Mice were initially anesthetized using a chamber containing gas anesthetic (oxygen,100%; isoflurane 4% for induction) and then positioned supine on a servo-controlled homeothermic pad (maintaining body temperature at 37°C). Anesthesia was maintained via a nose cone (100% O_2_ and 2% isoflurane). Right ventricle (RV) pressure-volume loop assessment was performed by invasive, closed-chest, right heart catheterization, on anesthetized mice, as described^36^. A high-fidelity catheter was advanced into the jugular vein and guided into the RV. Data were recorded continuously for 30-60 seconds using a Scisense ADV500 Pressure-Volume Measurement System (Transonic) and LabScribe2 software (iWorx, Dover, NH, USA). Right ventricular systolic and end diastolic pressure (RVSP, RVEDP) were measured. The mean pulmonary arterial pressure (mPAP) was calculated using the formula: mPAP=(0.61×RVSP+2) mmHg^37^. The catheter was then gently withdrawn and soaked in a heparin-sodium chloride solution. Later, a horizontal incision of the skin and the underlying muscles was performed just below the xiphoid process of the sternum. The diaphragm was blunt dissected, and the LV was perforated with a 25G needle. The catheter was then inserted into the LV, and we measured left ventricular systolic and end diastolic pressure (LVSP, LVEDP).

#### 2.4 Tissue and Cell Processing

Lung, RV, LV, spleen, and thymus tissues were harvested and prepared for immunohistochemistry, flow cytometry or western blotting.

#### 2.5 Immunohistochemistry

Paraffin sections (5 µm) of the lung and RV were deparaffinized and hydrated. Antigen retrieval was achieved by microwave heating using citrate buffer (10 mM, pH 6.0; 20 min). Sections were allowed to cool and washed with PBS-tween 0.05% (PBS-T). For immunofluorescence staining, sections were blocked with 1% Bovine Serum Albumin (BSA) for 30 min at room temperature. Sections were incubated overnight with primary antibody (CD45 and/or DNMT3A) at 4°C (**Supplemental methods**). Sections were washed with PBS-T and incubated with conjugated secondary antibody for 1-hour at room temperature (**Supplemental methods**). Slides were washed and mounted using ProLong Diamond Mounting with DAPI (ThermoFisher; Waltham, MA, USA). The fluorescence-based imaging was performed using TCS SP8 laser scanning confocal microscopy (SP8 Leica, Concord, ON, Canada) and the HC PL APO CS2 63x/1.40 oil objective. The quantification of CD45+ cells was performed using the LAS-X software (Leica) and is shown as total number of CD45+ cells in a field (0.02 mm^2^).

#### 2.6 Hematoxylin and Eosin (H&E)

Paraffin sections of lung were deparaffinized and hydrated. Sections were stained using hematoxylin and eosin (H&E Staining Kit ab245880, Abcam). Morphometric analysis was performed to measure pulmonary arterial wall thickness using ImageJ. The pulmonary artery medial wall thickness was estimated by measuring the outer and inner layers of arteries in transverse sections, and it was calculated as follows: [1-(perimeter of inner layer/perimeter of outer layer)] *100%.

#### 2.7 Collagen deposition quantification

Paraffin sections of RV tissue were deparaffinized and hydrated. The nuclei were stained with Wiegert’s hematoxylin and subsequently stained with Picrosirius Red Stain as per the manufacturer’s instruction (Sigma Aldrich; “Direct Red 80” Cat#365548). Images were acquired using a Leica DM 4000 microscope equipped with a DFC310 FX camera using a 20x objective and LASX software. The collagen fibers (red stain) were quantified using ImageJ by measuring the percentage red signal/field.

#### 2.8 Western Blotting

Total thymus and spleen protein was prepared in lysis, anti-protease buffer (#9803; Cell Signaling Technologies, Beverly, MA, United States) and measured using the BCA protein reagent assay (A/B) (Thermo scientific; cat# 23228, 1859078). 50µg (thymus) and 80 µg (spleen) of protein were loaded on a 4-12% gradient polyacrylamide gel for electrophoretic separation using the SDS-PAGE method. Running buffer was prepared using 20X MOPS Buffer (Novex: REF NP0001), 0.1% Bolt Antioxidant (Invitrogen: REF BT0005) and ddH_2_O. Resolved proteins were transferred to polyvinylidene fluoride (PVDF) membranes (Novex cat# LC2002) and blocked in 5% milk. Immunoblotting was performed using the primary antibodies as described in **Supplemental methods**. Membranes were developed using an ECL Western blot detection reagent (Amersham: REF RPN2232), imaged using ChemiDoc^TM^MP imaging system (Bio-Rad Laboratories; Mississauga, ON, Canada) and analyzed with ImageJ. β-actin was used as a loading control for data normalization.

#### 2.9 Flow cytometry

The lungs were harvested and kept in a calcium-free Hanks medium (Sigma Aldrich) until processing. The lungs were minced and digested using DNase (100 mg/mL) and Liberase (13 WU/mL) for 30 minutes at 37°C (2 cycles of 15 minutes). The cell suspensions of lung were filtered through a 70 µm cell strainer, and the total cell number was calculated (2 x 10^5^ cells per tube). The LIVE/DEAD™ Fixable Near-IR Dead Cell Stain Kit (ThermoFisher Scientific L10119) was used to discriminate dead cells, per the manufacturers’ recommendation. Cells were stained using fluorescent-tagged antibodies against surface antigens (**Supplemental methods**) and washed with FACS buffer. Cells were fixed with 2% PFA (15 minutes, 4°C). The SH-800S cell sorter/cytometer (Sony Biotechnology San Diego, CA, USA) was used to acquire the cytometric data. The data was analyzed using FlowJo^TM^ (BD Life Sciences, USA).

#### 2.10 Plasma Cytokine Analysis

Blood was drawn via cardiac puncture following heart catheterization, collected in heparinized blood collection tubes and centrifuged at 2,000 x g for 15 minutes (Thermo Fisher Scientific 21R microcentrifuge, Waltham, MA, USA) and plasma was collected. A mouse cytokine, 32-Plex Discovery Assay® (MilliporeSigma, Burlington, Massachusetts, USA) was performed using the Luminex™ 200 system (Luminex, Austin, TX, USA; performed by Eve Technologies Corp., Calgary, Alberta).

#### 2.10 Statistical Analysis

Experimental animal randomization was implemented as per Provencher et al. 2018^38^. Operators were blinded to genotype and normoxic/hypoxic conditions. Results are presented as mean±SEM using GraphPad Prism (GraphPad Software, La Jolla, CA). An ANOVA or Student’s *t*-test were used to assess intergroup differences. Data was analyzed for normality. Non-parametric testing was used for data that was not normally distributed. A *p-*value <0.05 was considered statistically significant. The sample size required to obtain 80% power was 5-7 mice/group.

#### 2.11 Sex

Mice of both sexes were used. In the absence of a sex-related difference in data, male and female values were pooled.

## Results

### 1- Rare Deleterious Predicted Germline and Somatic DNMT3A Variants Are Increased in PAH Biobank Patients and are Associated with Lack of Acute Vasodilator Response

1832 unrelated European PAH patients were initially selected from the PAH Biobank to be compared to 7509 non-Finnish European gnomAD controls that were age-and sex-matched. Deleterious variants identified in *DNMT3A* in PAH WES data were tested using gene-based, case-control association analysis. A REVEL score >0.5 was used to define missense (D-MIS) variants. We tested for association in two categories of variants: D-MIS or D-MIS and likely gene damaging (D-MIS+LGD). Among all PAH cases, we observed a trend towards the enrichment of D-MIS variants for *DNMT3A* (6/1832 cases vs. 8/7509 controls; RR=3.1, p=0.04), though this did not reach the Bonferroni-corrected threshold for significance (**Table 1A**). The association was primarily due to patients with APAH (RR=4.5, p=0.01) (**Table 1B**).

**Table 1A.** Enrichment of rare predicted deleterious variants* in candidate PAH risk gene, *DNMT3A*, among European PAH cases.

| *DNMT3A* ** |  |  |  |  |
| --- | --- | --- | --- | --- |
| **Mutation type\*\*\*** | **PAH cases (n=1832)** | **gnomAD WGS controls (n=7509)** | **Relative risk** | **P-value** |
| **D-MIS** | 6 | 8 | 3.1 | 0.04 |
| **D-MIS+LGD** | 7 | 10 | 2.86 | 0.06 |

**Table 1B.**
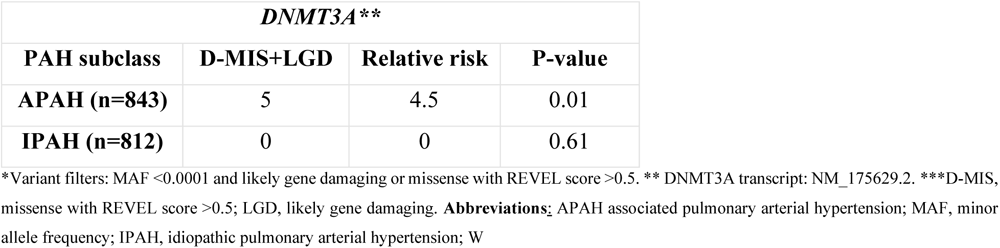
Enrichment of rare predicted deleterious variants* in candidate PAH risk gene, *DNMT3A*, among European APAH and IPAH cases.

| *DNMT3A* ** |  |  |  |
| --- | --- | --- | --- |
| **PAH subclass** | **D-MIS+LGD** | **Relative risk** | **P-value** |
| **APAH (n=843)** | 5 | 4.5 | 0.01 |
| **IPAH (n=812)** | 0 | 0 | 0.61 |

\*Variant filters: MAF <0.0001 and likely gene damaging or missense with REVEL score >0.5. \*\* *DNMT3A* transcript: NM\_175629.2. \*\*\*D-MIS, missense with REVEL score >0.5; LGD, likely gene damaging. **Abbreviations:** APAH associated pulmonary arterial hypertension; MAF, minor allele frequency; IPAH, idiopathic pulmonary arterial hypertension; W

Subsequently, sequencing data from the entire PAH (WES), gnomAD (WGS) and SPARK (WES) databases were analyzed for *DNMT3A* predicted deleterious variants. The prevalence of *DNMT3A* mutations was found to be significantly higher in the PAH Biobank [1% (27/2707 patients)] when compared to SPARK [0.33% (37/11198 patients); RR-3.019 (95% CI: 1.865-5.015); P<0.001] or gnomAD [0.43% (613/141456 patients); RR-2.302 (95% CI: 1.563-3.390); p=0.0002] databases **(Figures 1A and 1B).** The PAH Biobank cohort included 44% IPAH, 48% APAH, and 4% FPAH cases and none had other pathogenic/suspected pathogenic PAH gene mutations. Most patients had adult-onset PAH, with a typical ratio of 3.7:1 female to male. The genetically determined ancestries were 72% European, 12% Hispanic, 11% African, and smaller percentages of other lineages. The age of the cohorts was closely comparable between the gnomAD and PAH databases (**Figure 1C**).

We then analyzed the PAH Biobank cohort of 2572 cases. We identified 12 unique likely germline variants (4 LGD, 8 D-MIS) in 12 patients **(Supplemental S1**) and 17 unique likely somatic variants (3 LGD, 14 D-MIS) in 21 patients **(Table S2).** Two *DNMT3A* variants (p. Arg635Trp and c.1937-2A>G splicing) occurred as either likely germline or likely somatic mutations in different patients. There were three recurrent D-MIS variants, one recurrent splice variant, and one recurrent stop/gain variant for *DNMT3A*. Additionally, we observed variants in two CHIP mutation hotspots in *DNMT3A* (p.Arg882Cys and p.Arg882His)^39^ in five patients. None of the *DNMT3A* mutant patients had variants in known PAH risk genes^40–41^. However, subject 13-025, however, carried one *DNMT3A* splice variant and one *TET2* frameshift variant.

Two-dimensional structures of the predicted proteins showed that most of the DNMT3A missense variants (both predicted germline and somatic) localize to conserved protein domains, more than half of the patients (24/33) had gene variants predicted to affect the DNA methylation domain. None of the individuals with *DNMT3A* variants (0/19) were acute vasodilator responders, as assessed by standard criteria vs. 140/1043 (13.4%; p<0.05) acute vasodilator responders in the remainder of the PAH Biobank cohort **(Table S3).** Consistent with the severity associated with lack of vasodilator responsiveness, the use of endothelin receptor inhibitors and soluble guanylate cyclase stimulators was increased in individuals with *DNMT3A* variants versus the remainder of the PAH Biobank patients (p<0.05).

Two-thirds (21/33) of the patients with rare, predicted germline or somatic *DNMT3A* variants had APAH. The higher female: male ratio among predicted germline variant carriers (11:1) compared to the overall cohort likely reflects a large number of APAH patients with autoimmune diseases. Patients with *DNMT3A* variants and APAH exhibited trends toward increased age-of-onset and more moderate hemodynamic parameters; but these parameters were not significantly different from the overall APAH+IPAH cohort **(Tables S1 and S2)**.

To increase our sensitivity to detect somatic *DNMT3A* variants, targeted panel sequencing was performed to determine the prevalence of CHIP mutations in 710 PAH Biobank patients compared to 3645 individuals from a pooled control cohort. The prevalence of CHIP mutations was significantly higher in the PAH group (14.65%; 104/710) compared to controls after adjusting for participant age and sex (OR: 1.40 (95% CI: 1.09-1.80; p = 0.0071) (**Figures 2A**). *DNMT3A* was the most commonly mutated gene, accounting for almost half (49/104 = 47%) of mutations in PAH patients (**Figures 2B and C**). The trend for CHIP driven by *DNMT3A* alone (*DNMT3A-*CHIP) to be associated in PAH patients versus controls did not achieve statistical significance after adjusting for age and sex (OR 1.15, 95% CI: 0.82-1.63, p = 0.40; **Figure 2D**). Approximately 45% (22/49) of *DNMT3A* CHIP patients with PAH had APAH **(Table 2).** Hemodynamics were comparable between PAH patients with and without *DNMT3A* mutations **(Table 2).**

PAH patients were also screened for blood disorders, including anemia, neutropenia, thrombocytopenia, lymphocytopenia and homocysteinemia. Six *DNMT3A* CHIP patients were found to have anemia (PAH-05-002, PAH-05-158, PAH-05-068, PAH-05-097, PAH-08-037 and PAH-08-042) while one patient was identified with homocysteinemia (PAH-28-033). Several patients possessed mutations in *DNMT3A* hotspots. We identified three patients with *DNMT3A* mutations in Arg326 (two R326H and one R326C), three with mutations in Arg771 (all R771Q), and five with mutations in Arg882 (two R882C, two R882H and one R882S). CHIP driven by non-*DNMT3A* mutations (non-*DNMT3A* CHIP) was significantly enriched in the PAH group compared to controls (OR: 1.71, 95% CI: 1.23-2.38, p = 0.002, **Figure 2D**), and *TET2* was the most commonly mutated non-*DNMT3A* CHIP gene (18/104 = 17%, **Figures 2B and C**).

**Figure 2:**
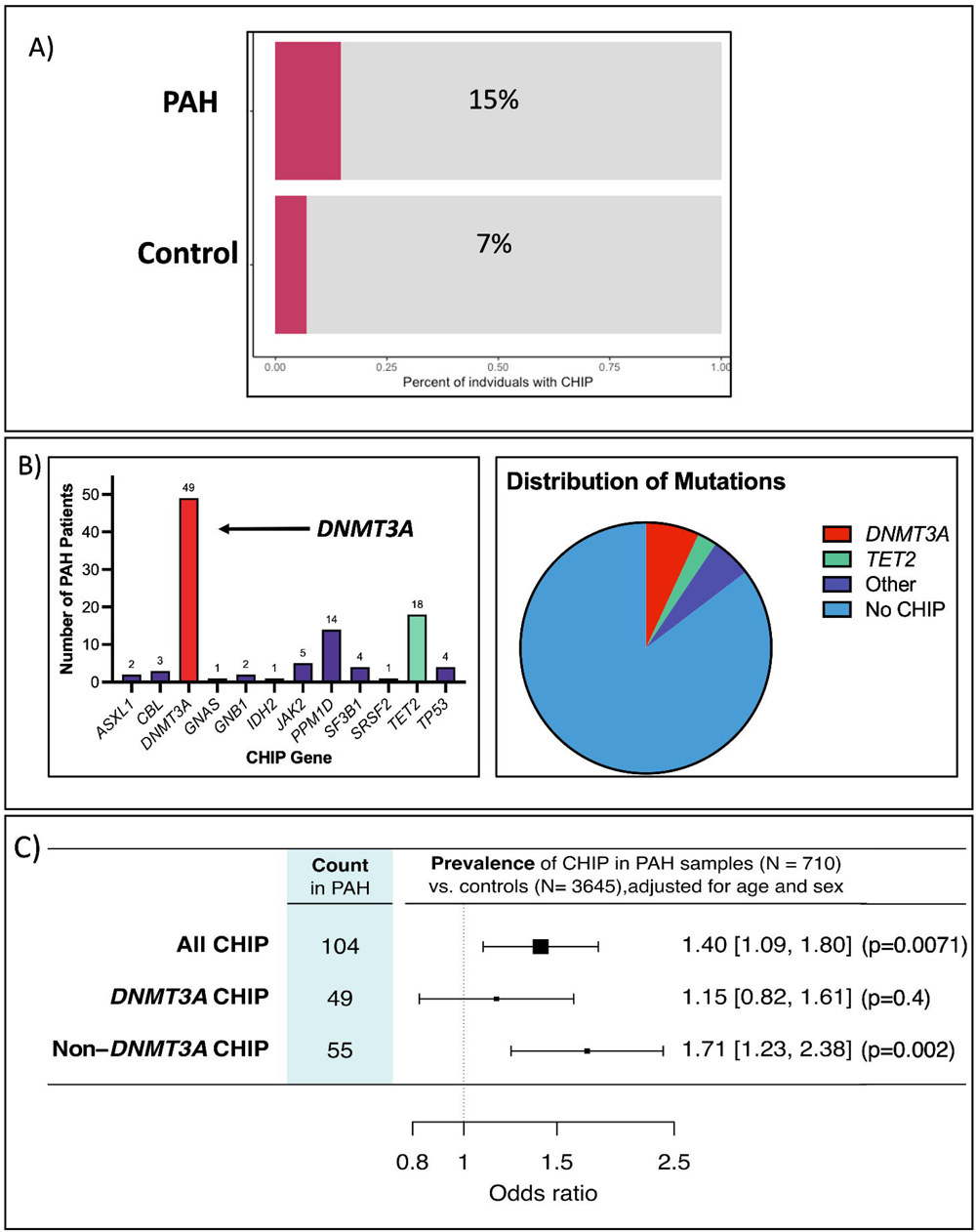
Prevalence of DNMT3A Mutations in PAH Patients using Targeted Panel Sequencing: A Comparative Analysis with a Pooled Control Population. **A)** Data was obtained using deep, targeted panel sequencing. The percentage of individuals with CHIP is shown for PAH patients (n=710) and controls (n=3645). Approximately 15% of PAH patients possessed a CHIP mutation while only 7% of controls were found to have CHIP. **B)** The number of patients with CHIP mutations in each gene is displayed. Number of patients is shown on the y-axis of the graph while genes are shown on the x-axis. *DNMT3A* was the most mutated gene (red; 49 patients), followed by *TET2* (green, 18 patients). All other mutated genes are shown in purple. The pie chart shows the mutation distribution using the same colours and displays the number of patients with no CHIP mutations in blue. **C)** The prevalence of all CHIP mutations, *DNMT3A* CHIP and non-*DNMT3A* CHIP is shown. 104/710 PAH patients were found to have CHIP mutations (49 being in *DNMT3A*, and 55 in other genes). The odds ratios, with 95% confidence intervals, are also shown for PAH patients with all CHIP mutations, *DNMT3A* CHIP and non-*DNMT3A* CHIP compared to controls. The odds ratios for all CHIP mutations and non-*DNMT3A* CHIP mutations were statistically significant (p=0.0071, p=0.002, respectively), though *DNMT3A* CHIP alone did not reach statistical significance (p=0.40).

**Table 2.** Rare, predicted deleterious likely somatic variants in candidate PAH risk gene, *DNMT3A*, among 710 PAH Biobank cases using targeted panel sequencing.

|  | Patient ID | Nucleotide change | Amino acid change | VAF | PAH class | Sex | Race* | Mean PAP (mmHg) | Mean PCWP, (mmHg) | CO, Fick (L/min) | PVR, Fick (Wood units) | Mean SAP (mmHg) | Mean SAP:PAP |
| --- | --- | --- | --- | --- | --- | --- | --- | --- | --- | --- | --- | --- | --- |
| 1 | 02-002 | c.G2312A | R771Q | 10% | APAH | F | White | 104 | 12 | NA | NA | 155 | 1.49 |
| 2 | 02-010 | c.1123delG | V375Wfs*32 | 3.5% | IPAH | F | White | 45 | 14 | NA | NA | 65 | 1.44 |
| 3 | 02-116 | c.A2204G | Y735C | 2.2% | IPAH | F | White | 47 | 5 | 3.52 | 11.93 | 75 | 1.60 |
| 4 | 03-061 | c.C976T | R326C | 10% | IPAH | M | Black | 57 | 12 | 6.94 | 6.48 | 87 | 1.53 |
| 5 | 04-041 | c.1063delC | H355Tfs*52 | 2.7% | IPAH | M | White | 84 | 6 | 4.73 | 16.49 | 132 | 1.57 |
| 6 | 5-002 | c.G1267T | E423X | 2.9% | IPAH | F | White | 56 | 13 | 5.41 | 7.95 | 79 | 1.41 |
| 7 | 05-026 | c.1148delT | L383Rfs*24 | 2.1% | IPAH | F | White | 66 | 2 | NA | NA | 112 | 1.70 |
| 8 | 05-034 | c.G977A | R326H | 6% | IPAH | F | White | 45 | 4 | 4.78 | 8.58 | 69 | 1.53 |
| 9 | 05-068 | c.1554+1G>A:c.987+1G>A:c.1554+1G>A:c.1098+1G>A | N/A | 7.5% | IPAH | F | White | 47 | 9 | NA | NA | 82 | 1.74 |
| 10 | 05-083 | c.G2312A | R771Q | 5.6% | IPAH | F | White | 47 | 14 | 3.02 | 10.93 | 70 | 1.49 |
| 11 | 05-097 | c.945_946insGATG | T316Dfs*9 | 2.6% | APAH | F | White | 39 | 4 | 3.98 | 8.79 | 58 | 1.49 |
| 12 | 05-101 | c.2082+1G>T:c.1515+1G>T:c.2082+1G>T:c.1626+1G>T | N/A | 4.5% | APAH | F | White | 31 | 8 | 5.98 | 3.85 | 49 | 1.58 |
| 13 | 05-134 | c.C2644T | R882C | 5% | IPAH | F | White | 36 | 13 | 3.81 | 6.04 | 50 | 1.39 |
| 14 | 05-145 | c.G1136A | R379H | 6.9% | APAH | F | White | 43 | 7 | NA | NA | 65 | 1.51 |
| 15 | 05-158 | c.T1220G | I407S | 2.3% | IPAH | F | Black | 59 | 13 | 3.48 | 13.22 | 83 | 1.41 |
| 16 | 05-185 | c.G2312A | R771Q | 9.6% | APAH | F | White | 47 | 15 | 4.01 | 7.98 | 81 | 1.72 |
| 17 | 05-202 | c.2174-2A>G:c.1607-2A>G:c.2174-2A>G:c.1718-2A>G | N/A | 26.4% | IPAH | F | White | 35 | 14 | 6.76 | 3.11 | 47 | 1.34 |
| 18 | 06-074 | c.1279+1G>A:c.712+1G>A:c.1279+1G>A:c.823+1G>A | N/A | 3.6% | IPAH | F | White | 33 | 7 | 3.2 | 8.13 | 49 | 1.48 |
| 19 | 07-051 | c.C1671A | C557X | 2% | APAH | F | White | 47 | 10 | 4 | 9.25 | 80 | 1.70 |
| 20 | 07-099 | c.C1560A | C520X | 3.5% | IPAH | F | Asian | 53 | 13 | 2.23 | 17.94 | 80 | 1.51 |
| 21 | 08-037 | c.G1906A | V636M | 2% | IPAH | F | White | 50 | 3 | NA | NA | 88 | 1.76 |
| 22 | 08-042 | c.A1979G | Y660C | 9.5% | IPAH | F | White | 45 | 6 | 2.3 | 16.96 | 76 | 1.69 |
| 23 | 08-066 | c.A1894T | K632X | 2% | APAH | M | White | 65 | 13 | 4.8 | 10.83 | 118 | 1.82 |
| 24 | 10-028 | c.C1543T | Q515X | 2.3% | IPAH | M | White | 46 | 3 | NA | NA | 80 | 1.74 |

|  | Patient ID | Nucleotide change | Amino acid change | VAF | PAH class | Sex | Race* | Mean PAP (mmHg) | Mean PCWP, (mmHg) | CO, Fick (L/min | PVR, Fick (Wood units) | Mean SAP (mmHg) | Mean SAP:PAP |
| --- | --- | --- | --- | --- | --- | --- | --- | --- | --- | --- | --- | --- | --- |
| 25 | 10-060 | c.C2193A | F731L | 4.1% | IPAH | M | White | 63 | 9 | 2.99 | 18.06 | 91 | 1.44 |
| 26 | 10-063 | c.C1816T | Q606X | 3% | APAH | F | White | 61 | 7 | 3.12 | 17.31 | 94 | 1.54 |
| 27 | 12-036 | c.G977A | R326H | 3% | IPAH | F | White | 49 | 6 | NA | NA | 89 | 1.82 |
| 28 | 12-051 | c.2408+1G>C:c.1841+1G>C:c.408+1G>C:c.1952+1G>C | N/A | 2.9% | IPAH | F | White | 58 | 9 | NA | NA | 95 | 1.64 |
| 29 | 12-057 | c.G1969A | V657M | 9.1% | APAH | F | White | 51 | 3 | 4.16 | 11.54 | 83 | 1.63 |
| 30 | 12-114 | c.T2114C | I705T | 3% | IPAH | F | White | 35 | 13 | 3.83 | 5.74 | 49 | 1.40 |
| 31 | 12-173 | c.G2645A | R882H | 10.8% | APAH | M | White | 55 | 16 | NA | NA | 83 | 1.51 |
| 32 | 13-021 | c.G2645A | R882H | 16% | APAH | F | White | 35 | 11 | 6.9 | 3.48 | 58 | 1.66 |
| 33 | 13-027 | c.C2644A | R882S | 2.3% | APAH | F | White | 25 | 8 | 4.8 | 3.54 | 42 | 1.68 |
| 34 | 13-028 | c.C2329A | P777T | 5.4% | APAH | M | White | 45 | 8 | NA | NA | 70 | 1.56 |
| 35 | 14-002 | c.978_985del | W327Vfs*13 | 3.5% | APAH | F | White | 37 | 13 | NA | NA | 54 | 1.46 |
| 36 | 17-004 | c.1937-2A>G:c.1370-2A>G:c.1937-2A>G:c.1481-2A>G | N/A | 8.6% | APAH | F | White | 28 | 7 | 7.17 | 2.93 | 48 | 1.71 |
| 37 | 19-024 | c.C920G | P307R | 5.9% | IPAH | F | White | 36 | 10 | 5.91 | 4.40 | 56 | 1.56 |
| 38 | 19-040 | c.2252_2255del | F751Sfs*27 | 11.8% | IPAH | M | White | 54 | 9 | 5.48 | 8.21 | 90 | 1.67 |
| 39 | 21-001 | c.G2386T | G796C | 2.7% | IPAH | F | Black | 38 | 10 | 5.3 | 5.28 | 60 | 1.58 |
| 40 | 22-013 | c.C2206T | R736C | 12.4% | APAH | F | White | 32 | 8 | 7.1 | 3.38 | 53 | 1.66 |
| 41 | 22-052 | c.A2281G | M761V | 4.7% | IPAH | F | White | 60 | 3 | 3.53 | 16.15 | 100 | 1.67 |
| 42 | 22-058 | c.T2724G | Y908X | 8.2% | APAH | F | White | 33 | 8 | NA | NA | 50 | 1.52 |
| 43 | 22-126 | c.1667+1G>A:c.1100+1G>A:c.667+1G>A:c.1211+1G>A | N/A | 4.1% | APAH | F | White | 42 | 17 | 3.3 | 7.58 | 68 | 1.62 |
| 44 | 28-033 | c.C1096T | R366C | 14.5% | IPAH | F | White | 61 |  | 3.6 | NA | 89 | 1.46 |
| 45 | 29-026 | c.T2194A | F732I | 12% | APAH | F | Black | 64 | 8 | 3.17 | 17.67 | 100 | 1.56 |
| 46 | 29-037 | c.C2644T | R882C | 8.9% | APAH | F | White | 88 | 16 | 3.87 | 18.60 | 125 | 1.42 |
| 47 | 29-041 | c.846dupA | E283Rfs*41 | 2.1% | APAH | F | Black | 27 | 11 | NA | NA | 43 | 1.59 |
| 48 | 29-060 | c.G2259A | W753X | 6.2% | APAH | F | Black | 63 | 11 | 3.72 | 13.98 | 106 | 1.68 |
| 49 | 35-014 | c.C2227T | P743S | 3.2% | IPAH | F | White | 49 | 13 | NA | NA | 110 | 2.24 |
| Mean ± SD, DNMT3A carriers |  |  |  |  |  | 5.13:1 |  | 49 ± 16 | 9.5 ± 3.9 | 4.4 ± 1.4 | 9.9 ± 5.2 | 78 ± 25 | 1.6 ± 0.2 |
| n, DNMT3A carriers |  |  |  |  |  |  |  | 49 | 48 | 34 | 33 | 49 | 49 |

\**DNMT3A* transcript: NM\_175629.2. Data obtained via deep, targeted panel sequencing of 710 PAH Biobank Patients. **Abbreviations:** APAH, associated pulmonary arterial hypertension; FPAH, familial pulmonary arterial hypertension; IPAH, idiopathic pulmonary arterial hypertension; PAP, pulmonary artery pressure; PCWP, pulmonary capillary wedge pressure; PVR, pulmonary vascular resistance; SAP, systemic arterial pressure. \* = self-reported

### 2- DNMT3A Expression is Reduced in Peripheral Blood Mononuclear Cells (PBMCs) of PAH Patients

We next investigated *DNMT3A* expression in peripheral blood mononuclear cells (PBMCs) in a separate patient cohort. Gene expression omnibus (GEO) analysis was conducted on samples from 50 SSc-PAH patients, 30 IPAH patients, 19 scleroderma patients without PAH (SSc), and 41 healthy controls. All cases had adult-onset PAH, with genetically determined ancestries as follows: 82.1% Caucasian, 14.3% African, and smaller percentages of other ancestries. The sex ratios (female: male) were as follows: control 4.9:1, IPAH 5:1, SSc-PAH 3.7:1, cohort characteristics previously published by *Potus et al*. (2020)^20^. We examined the expression of DNMT3A transcript variants 2 (NM_153759.3), 3 (NM_022552), and 4 (NM_175630.1), which code for the three protein isoforms DNMT3A2, DNMT3A3, and DNMT3A4, respectively (**Figure 3A**). Our analysis revealed decreased expression of variant 4 in 88% of SSc-PAH and 93.3% of IPAH patients (IPAH 0.77; SSc-PAH 0.8; p<0.0001), as well as decreased expression of variant 3 in 80% of SSc-PAH and 73.3% of IPAH patients compared to healthy individuals (IPAH 0.92; SSc-PAH 0.93; p<0.05) (**Figures 3B and C**). However, the gene expression of variant 2 was unaffected (**Figure 3D**). Notably expression levels were not altered in SSc patients without PAH versus control, indicating that the depressed expression is related to PAH not to scleroderma alone. To further assess the potential of DNMT3A variants 3 and 4 as predictors of PAH, we performed ROC analysis on a cohort consisting of 41 healthy controls and 80 PAH patients (IPAH/SSc-PAH). DNMT3A variant 4 (area under curve, AUC: 0.82; p<0.0001) and variant 3 (AUC: 0.67; p<0.002) could serve as potential predictors of PAH (**Figure 3E-G**).

**Figure 3:**
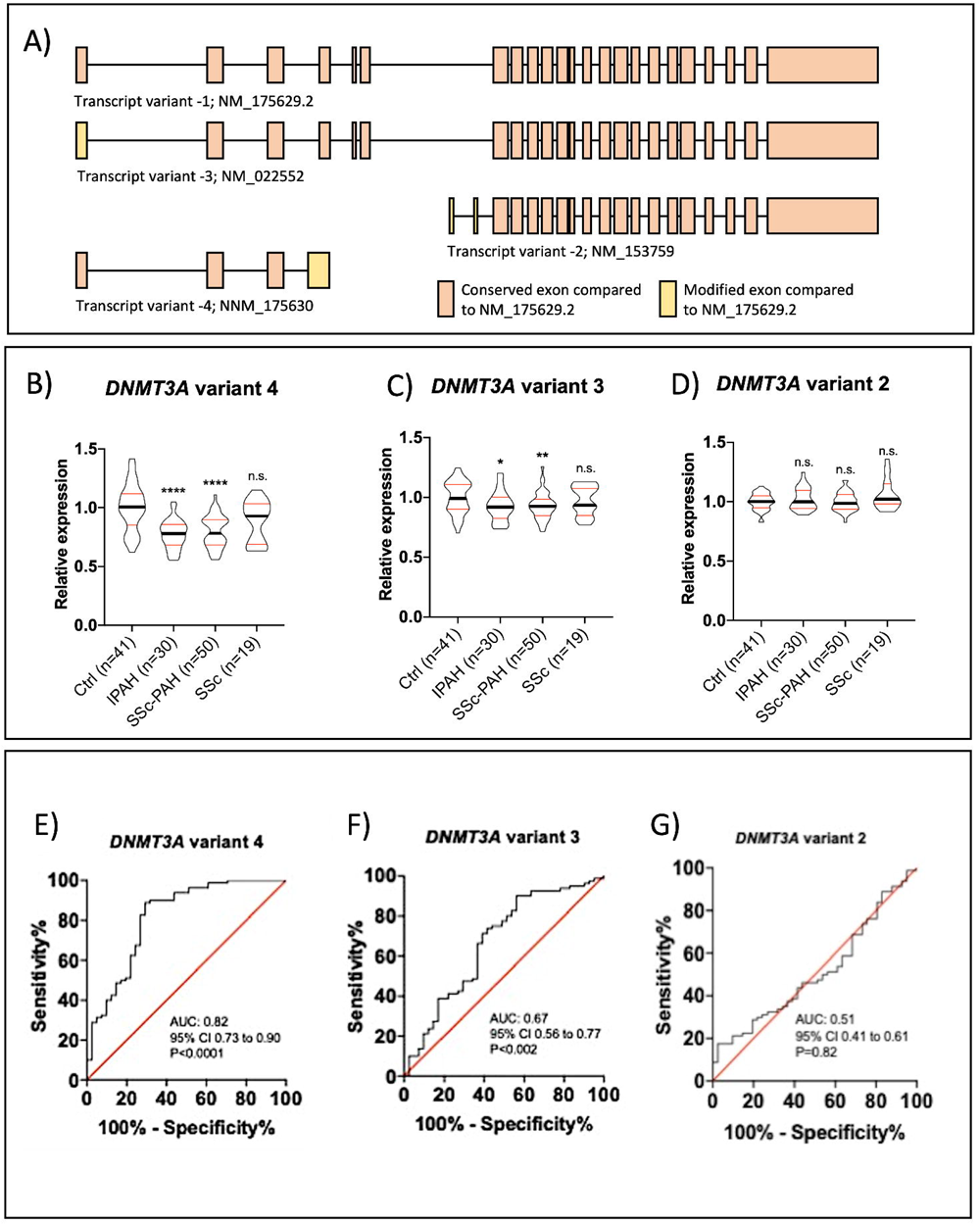
*DNMT3A* Expression is Reduced in Peripheral Blood Mononuclear Cells (PBMCs) of PAH Patients. **A)** The three DNMT3A transcript variants 2 (NM_153759.3), 3 (NM_022552), and 4 (NM_175630.1), which code for the three protein isoforms DNMT3A2, DNMT3A3, and DNMT3A4, respectively, are shown, in comparison to the original DNMT3A transcript variant 1. **B-D)** Gene expression of the DNMT3A isoforms 2,3 and 4 was measured in PBMCs. Decreased expression of variant 4 in 88% of SSc-PAH and 93.3% of IPAH patients (IPAH 0.77; SSc-PAH 0.8; p<0.0001), as well as decreased expression of variant 3 in 80% of SSc-PAH and 73.3% of IPAH patients compared to healthy individuals (IPAH 0.92; SSc-PAH 0.93; p<0.05) was found. The gene expression of variant 2 was unaffected. **E-G)** Receiver operating characteristic (ROC) analysis on a cohort consisting of 41 healthy controls and 80 PAH patients (IPAH/SSc-PAH) was performed for DNMT3A isoforms 2, 3 and 4. Sensitivity % is shown on the y-axis while the x-axis represents 100% subtract the Specificity %. The results indicate that DNMT3A variant 4 (AUC: 0.82; p<0.0001) and variant 3 (AUC: 0.67; p<0.002) could serve as potential predictors of PAH.

### 3- Dnmt3a in the Immune Organs of Dnmt3a^f/f^ Mice is Eliminated in Hematopoietic Dnmt3a^-/-^ Mice

PCR genotyping of PBMCs and ear tissue shows the presence of the floxed *Dnmt3a* band (330 bp) in both the ear and PBMCs in control mice. The floxed *Dnmt3a* band is present in the ear but absent in the PBMCs of the knockout mice (-/-) **(Figure S1A),** confirming the hematopoietic-specific knockout (n=3/group). The Vav-iCre (bottom of gel; lower band - 236 bp) was only present in the knockout mice, compared to the internal control (upper band - 324 bp), demonstrating that the Cre-mediated *Dnmt3a* excision mechanism was only present in the knockout mice. At the protein level, Dnmt3a was measured in immune organs (thymus and spleen) in *Dnmt3a^f/f^* versus *Dnmt3a*^-/-^ mice. Expression of Dnmt3a in the thymus was also investigated using confocal microscopy. Dnmt3a was present in the CD45^+^ leukocytes of the *Dnmt3a ^f/f^* mice and absent in the CD45^+^ leukocytes of the *Dnmt3a^-/-^*mice **(Figure S1B**). Western blotting of spleen and thymus tissue revealed significantly higher expression of Dnmt3a in *Dnmt3a^f/f^* mice compared to the *Dnmt3a^-/-^* mice, in which protein was almost absent (Spleen: p=0.012, Thymus: p=0.027**, Figure S1C)** (n=3-4/group). ß-actin was used as the internal protein control.

### 4- Dnmt3a Depletion in Murine Hematopoietic Cells Promotes PAH Development and Induces RVF

Compared to control mice, 9-month-old *Dnmt3a*^-/-^ mice exhibited a significant rise in RVSP (p=0.0004) and mPAP (p=0.0004; **Figure 4A**). Additionally, there was a noticeable tendency towards increased RVEDP (**Figure 4A**). PAAT, TAPSE, and CO were all significantly diminished in comparison to *Dnmt3a^f/f^* controls (p=0.0003, p=0.012, p=0.006), compatible with RV failure (**Figure 4A**). Even at age 4.5 months, mice spontaneously developed PAH evident by increased RVSP (p=0.0015), low PAAT and TAPSE (p=0.0030, p=0.0001) and a trend of reduction in CO (p=0.2061). There was a trend to exacerbation of PAH by transient exposure to hypoxia for 3 weeks followed by a 3-week recovery period pre-cardiac catheterization (p=NS). 3-month-old *Dnmt3a^-/-^* mice exposed to chronic hypoxia developed PAH, characterized by elevated RVSP, mPAP, and RVEDP (p=0.0004, p=0.0007, p=0.0724) as well as reduced PAAT and TAPSE (p=0.0020, p=0.0222) compared to hypoxic *Dnmt3a^f/f^* mice (**Figure 4B**). CO data showed a trend of reduction in *Dnmt3a^-/-^* mice, but the difference was not significant. No significant differences were observed in the hemodynamic changes of the LV when comparing *Dnmt3a^-/-^* mice to controls, as indicated by normal LVSP, LVEDP, and mitral annular plane systolic excursion, MAPSE **(Figure S2).**

**Figure 4:**
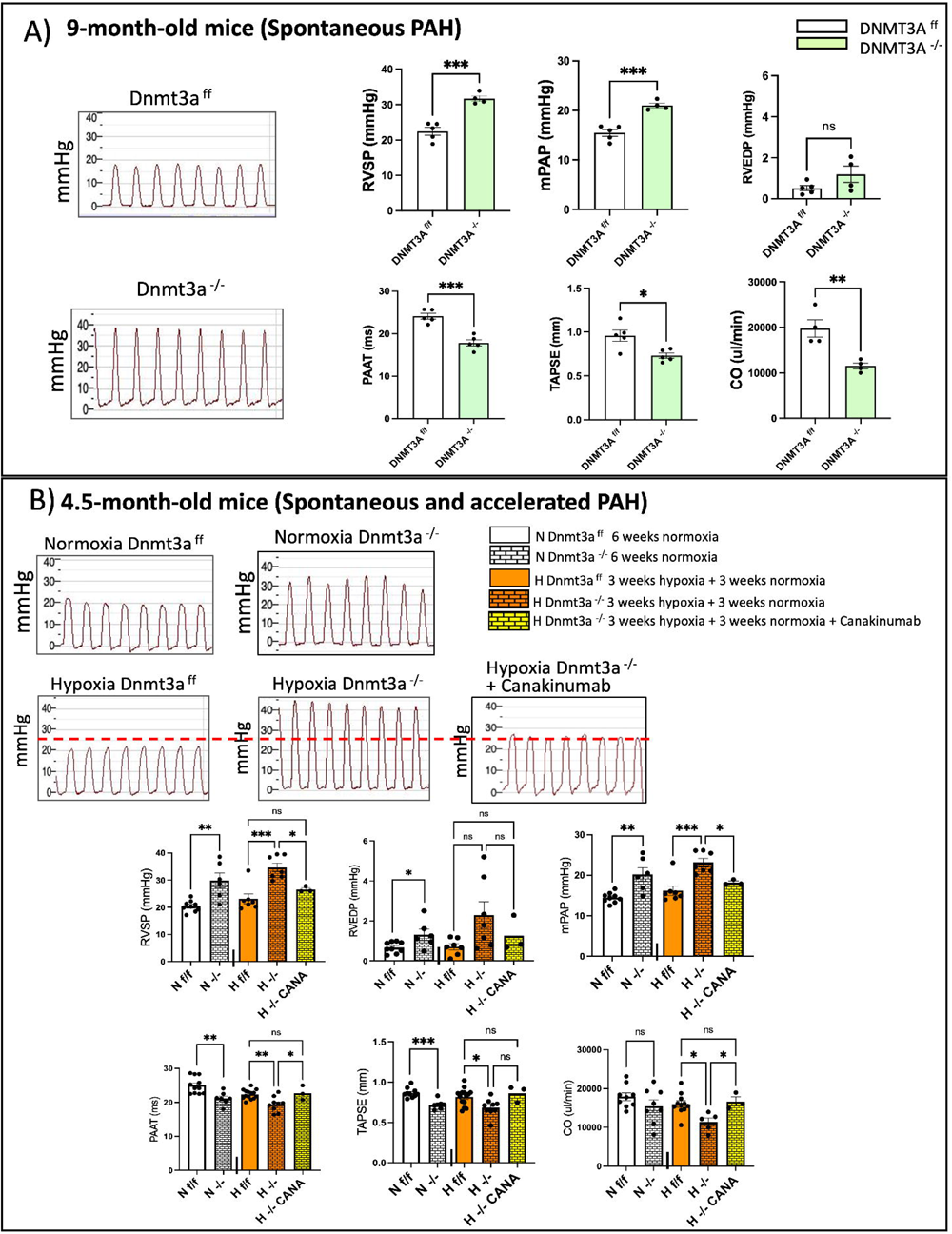
Dnmt3a Depletion in Mice HSCs Promotes PAH Development and Induces RVF, which can be Attenuated with Canakinumab. **A)** Hematopoietic *Dnmt3a*-knockout mice spontaneously develop PAH by 9 months of age (n=4-5/group). Right ventricular systolic pressure (RVSP; in mmHg) and mean pulmonary arterial pressure (mPAP; in mmHg) are significantly increased in *Dnmt3a*-knockout mice (green) compared to controls (white; p=0.0004, p=0.0004, respectively). Pulmonary artery acceleration time (PAAT; in ms), tricuspid annular plane systolic excursion (TAPSE; in mm) and cardiac output (CO; in ul/min) are significantly reduced in *Dnmt3a*-knockout mice compared to controls (p=0.0003, p=0.012; p=0.006, respectively). Right, ventricular end-diastolic pressure (RVEDP; in mmHg) shows a trend towards being increased in *Dnmt3a*-knockout mice, though not statistically significant. Right-heart catheterization (RHC) traces of RVSP are shown on the left for both control (top) and knockout (bottom) mice. RVSP, mPAP, and RVEDP were obtained via RHC, while PAAT, TAPSE and CO were obtained via echocardiography. **B)** Hematopoietic *Dnmt3a*-knockout mice develop PAH, accelerated by a second hit hypoxia (n=6-9/group). 3-month-old control (white boxes) and *Dnmt3a*-knockout (orange boxes) mice were either maintained in normoxic (solid colour boxes) conditions for 6 weeks or were exposed to 3 weeks of hypoxia followed by 3 weeks of normoxia (boxes with bricks). Representative RVSP traces obtained via RHC are shown for each group. RVSP, mPAP and RVEDP (trending) are elevated in *Dnmt3a*-knockout mice that underwent a second hit (hypoxia) compared to hypoxic controls (p=0.0004, p=0.0007, p=0.0724, respectively), and RVSP, mPAP and RVEDP are also significantly increased in *Dnmt3a*-knockout mice that were kept in normoxia compared to the normoxic controls (p=0.0015, p=0.0015, p=0.0234, respectively). PAAT and TAPSE are significantly reduced in *Dnmt3a*-knockout mice that underwent a second hit (hypoxia) compared to controls (p=0.0020, p=0.0222, respectively) and are also significantly increased in *Dnmt3a*-knockout mice that were kept in normoxia, compared to the normoxic controls (PAAT: p=0.0030; TAPSE: p=0.0001). CO was significantly reduced in hypoxic *Dnmt3a*-knockout mice compared to controls (p=0.0101), while normoxic *Dnmt3a*-knockout mice showed a trend towards being reduced, though not statistically significant (p=0.2061). * = p< 0.05. Treatment of hypoxic *Dnmt3a*-knockout mice with Canakinumab (yellow bars with bricks) improved hemodynamic measurements and cardiac function (n=3; RVSP: p=0.0362, mPAP: p=0.0449, PAAT: p=0.0258, TAPSE: p=0.0533, CO: p=0.0299). RVEDP was not significantly reduced (p=0.4629).

### 5- Hematopoietic Dnmt3a Depletion in Mice Induces Pulmonary Vascular Remodeling and RV Fibrosis

The hemodynamic changes observed in the 9-month-old *Dnmt3a^-/-^*mice were accompanied by adverse pulmonary vascular remodeling of small pulmonary arteries (diameter <50µm). Specifically, the pulmonary artery % medial wall thickness (PMWT) exhibited a significant increase compared to the control group (p=0.0023, **Figure 5A**). Exposure of 3-month-old *Dnmt3a^-/-^*mice to hypoxia exacerbated this adverse pulmonary vascular remodeling (P< 0.0001; **Figure 5B**). Also, there was an increase in total collagen deposition in the RV tissue of both normoxic and hypoxia accelerated *Dnmt3a^-/-^* mice indicated by the intensified Picrosirius red staining of RVs, when compared to *Dnmt3a^f/f^* controls (**Figure 5C**).

**Figure 5:**
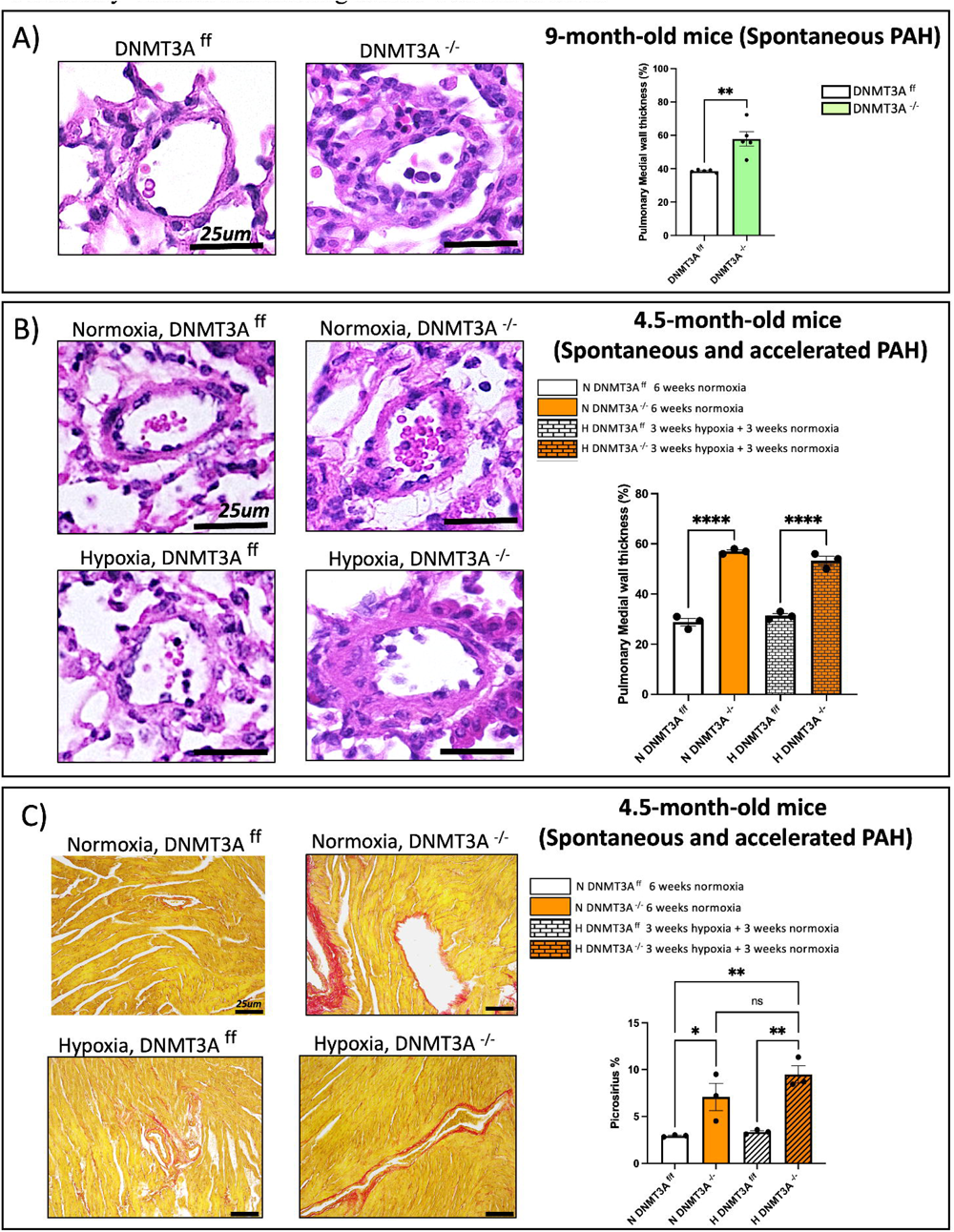
Dnmt3a depletion in HSCs induces pulmonary vascular remodeling and RV fibrosis in PAH. Histological assessment via hematoxylin and eosin (H&E) staining was performed on lung tissue slides and small pulmonary arteries were identified. Pulmonary medial wall thickness was compared between control and knockout mice. **A)** Pulmonary vascular remodelling occurs in the lungs of hematopoietic *Dnmt3a*-knockout mice by 9 months of age. There is a significant increase in the pulmonary medial wall thickness, defined as the % of the wall consisting of tunica media, in hematopoietic *Dnmt3a*-knockout mice (green) compared to controls (white; p=0.0005; n=5/group). A t-test was used to assess the difference between control and knockout mice. **(B)** There is an increase in pulmonary medial wall thickness in the lungs of hematopoietic *Dnmt3a*-knockout mice exposed to a second-hit hypoxia compared to controls (p<0.0001; n=3/group). 4.5-month-old *Dnmt3a*-knockout mice also spontaneously developed pulmonary vascular remodeling (p<0.0001; n=3/group). Bars with bricks represent hypoxic exposure while solid bars represent normoxia only. White represents control mice and knockout mice values are shown in orange. **(C)** Collagen deposition assessment of RV tissue using Picrosirius red staining (n=3/ group) demonstrated increased collagen deposition in the RV tissue of both hypoxic and normoxic *Dnmt3a*-knockout mice compared to their respective controls.

### 6- Hematopoietic Dnmt3a Depletion in Mice Leads to Pulmonary Leukocyte Accumulation, Dominated by Macrophages, and is Associated with Increased Plasma Levels of IL-13

Quantification of hemopoietic lineage cells (CD45+ cells) via confocal microscopy showed a significant increase in leukocytes in the lungs of 9-month-old *Dnmt3a^-/-^* mice versus *Dnmt3a^f/f^* controls (p=0.002, **Figure 6A**). Furthermore, 4.5-month-old *Dnmt3a^-/-^* mice also had increased pulmonary leukocyte infiltration (p=0.0473), though there was not a significant increase in CD45^+^ cells in the lungs of *Dnmt3a^-/-^* mice exposed to hypoxia compared to hypoxic *Dnmt3a^f/f^* controls (**Figure 6B**). Flow cytometric analysis of live lung leukocytes (CD45^+^ cells) collected from lung-derived, single-cell suspensions of normoxic *Dnmt3a^f/f^* and *Dnmt3a^-/-^* mice (n=5-8/ group) revealed increased leukocyte infiltration in the *Dnmt3a^-/-^* lungs (p=0.05) with macrophages (F4/80) being the most dominant cell type (p<0.01; **Figure 6C**). There was a trend towards an increase in neutrophils (LY6G; p=0.08), no noticeable difference in B-cells (CD19; p=0.46) and a decrease in T-cells (CD3; p=0.03) in the *Dnmt3a^-/-^* lungs (**Figure S3A**). Interleukin-13 (IL-13) was increased in the plasma of *Dnmt3a^-/-^*mice compared to controls (p=0.05; **Figure 6D**). There was also a trend towards an increase in granulocyte-colony stimulating factor (G-CSF) in *Dnmt3a^-/-^* mice plasma (p=0.16, **Figure 6D**).

**Figure 6:**
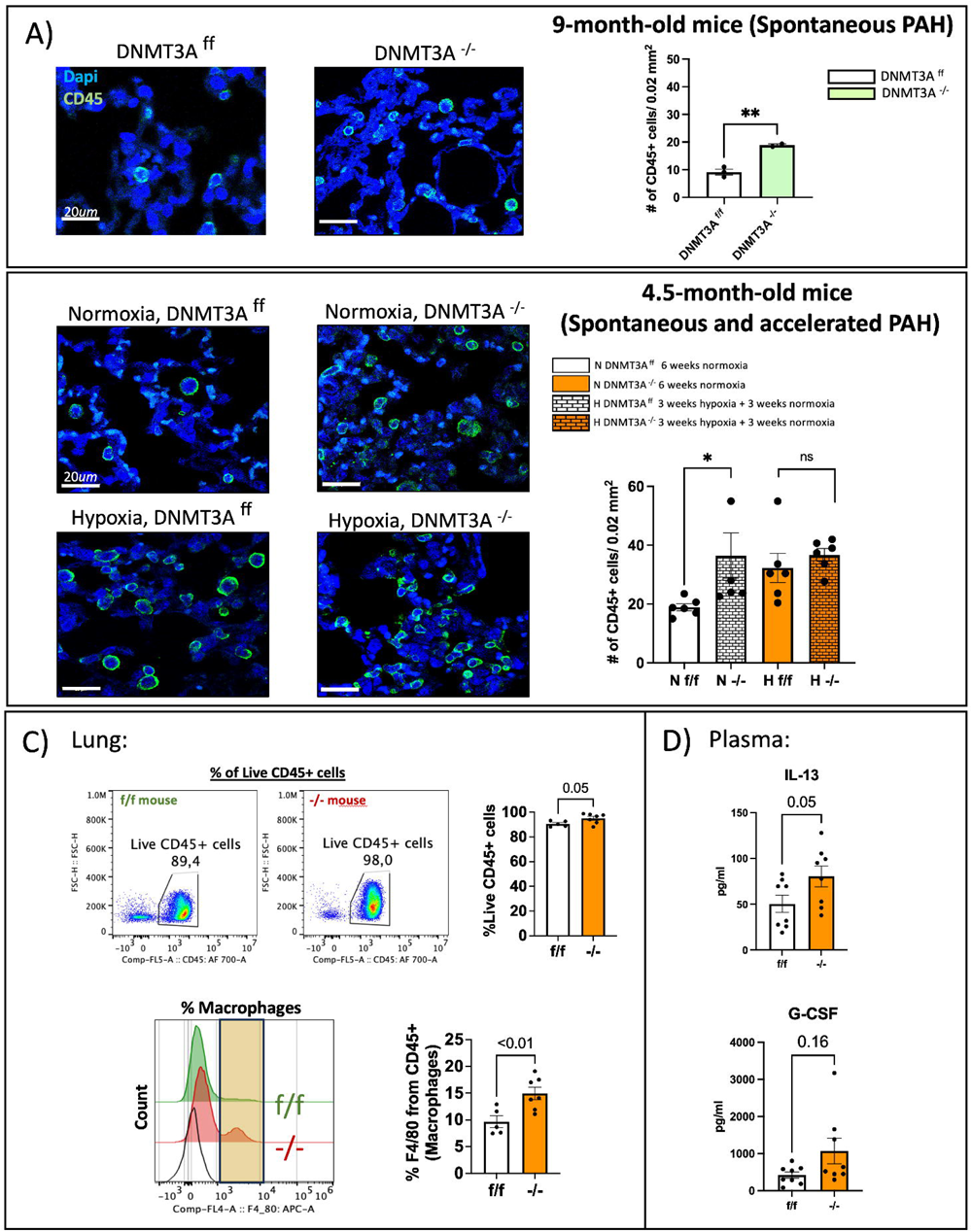
Dnmt3a Depletion in Mice HSCs Leads to Pulmonary Leukocyte Accumulation, Dominated by Macrophages, and is Associated with Increased Plasma Levels of IL-13. Immunofluorescence and confocal microscopy were used to measure leukocyte infiltration in the lungs of *Dnmt3a*-knockout mice with PAH compared to controls. CD45, a marker of leukocytes, is shown in green. DAPI, a nuclei stain, is shown in blue. **(A)** There is a trend towards an increase in the number of CD45+ cells in the lungs of nine-month-old knockout mice (green), that have developed PAH, compared to controls (white; n=2/group). The graph represents the number of CD45+ cells per 0.02 mm^2^. **(B)** There is a significant increase in the number of CD45+ cells in the lungs of the knockout mice, that have developed PAH, compared to controls (n=5/group, p=0.0473). Hypoxia appears to exacerbate leukocyte infiltration of the lungs, though there was no significant difference between knockout and control mice in hypoxia. Bars with bricks represent 3 weeks of hypoxic exposure with 3 weeks of normoxic exposure while solid bars represent 6 weeks of normoxia. White bars represent control mice and orange bars represent *Dnmt3a*-knockout mice. **(C)** Flow cytometry was used to investigate the subpopulations of leukocytes seen on confocal microscopy. Following a live/dead stain, CD45+ cells (leukocytes) were gated for and were found to be increased on 4.5-month-old *Dnmt3a*-knockout mice compared to controls (n=5-8/group; p=0.05). Specifically, macrophages are increased in *Dnmt3a*-knockout mice (n=5-8/group; p=<0.01). An LY6G antibody was used for neutrophil detection, an F4/80 antibody was used for macrophage detection, a CD3 antibody was used for T-cell detection, and a CD19 antibody was used for B-cell detection. On average, approximately 90% of CD45+ cells in the control mice were macrophages, while this was increased to approximately 95% in the *Dnmt3a*-knockout mice. **D)** IL-13 was found to be increased in the plasma of *Dnmt3a*-knockout mice compared to controls (p=0.05). There was also a trend towards an increase in plasma G-CSF in the *Dnmt3a*-knockout mice compared to controls (p=0.16). Plasma cytokine concentration is shown on the y-axis in picogram per milliliter (pg/ml).

### 7- Treatment with the IL-1β antagonist, Canakinumab, Improves Hemodynamics and Attenuates PAH in Dnmt3a^-/-^ Mice

Hypoxic *Dnmt3a^-/-^* mice treated with canakinumab had reduced RVSP and mPAP (p=0.0362, p=0.0449), as well as improved PAAT and CO (p=0.0258, p=0.0299) and a trend towards increased TAPSE (p=0.0533; **Figure 4B**). There were no significant differences in LVSP or LVEDP between groups. Mice treated with canakinumab were improved versus knockout mice and had hemodynamic measures comparable to hypoxic *Dnmt3a^f/f^* mice (RVSP: p=0.4731, mPAP: p=0.5278, PAAT: p=0.9437, TAPSE: p=0.7894; **Figure 4B**).

## Discussion

DNMT3A is an enzyme that mediates *de novo* gene methylation and is a well-established epigenetic regulator of gene expression^40^. Our study aimed to investigate *DNMT3A* as a potential PAH gene and explore the impact of impaired DNMT3A function on PAH development, considering the established role of epigenetic dysregulation in PAH etiology. This study revealed several noteworthy findings.

First, we found that *DNMT3A* variants were significantly more common in 1832 European PAH Biobank patients when compared to 7509 age and sex matched European individuals within the gnomAD database. When comparing all patients in each database, not age or sex matched, deleterious *DNMT3A* variants were also found to be significantly higher in PAH Biobank patients compared to participants in both gnomAD and SPARK control databases. We then used WES data from the PAH Biobank and identified significant enrichment of rare predicted deleterious germline variants in *DNMT3A*, along with additional likely somatic mutations. The majority of the cases with a *DNMT3A* mutation had APAH (21/33).

Second, deeper, targeted panel sequencing of 710 PAH Biobank patients and 3645 controls revealed a statistically significant association between PAH and somatic CHIP mutations. Though *DNMT3A* CHIP alone was not significant compared to controls, almost half of the PAH patients with CHIP mutations had mutations in *DNMT3A* (49/104) and approximately 45% (22/49) of them had APAH. These results suggest that DNMT3A dysfunction appears to be particularly important in APAH. The novelty of this finding is increased because these mutations notably occurred in patients who were free from other known PAH-associated gene variants, though only coding regions were analyzed. These data are the first report of genetic factors associated with APAH, beyond congenital heart disease.

Third, in an independent cohort studying DNMT3A expression in PBMCs, we observed a significant decrease in *DNMT3A* expression in IPAH and APAH patients. Notably, our ROC analysis demonstrated the potential of reduced DNMT3A expression to serve as a predictive biomarker for PAH. Although certain *DNMT3A* germline and CHIP variants may lead to decreased DNMT3A mRNA or protein expression^42^, the prevalence of decreased DNMT3A expression (in 73.3% to 93.3% of these PAH patients) far surpasses the prevalence of CHIP we observed (6.9%), suggesting other processes may be associated with decreased DNMT3A expression in PAH, signifying both its relevance to pathophysiology and its potential to serve as a biomarker.

Fourth, we utilized a mouse model with a targeted deletion of *Dnmt3a* in HSCs. This approach aimed to mimic the findings observed in human PBMCs with reduced DNMT3A expression and allowed study of the contribution of pathological DNMT3A depletion to the development of PAH. The mouse data provided compelling biologic plausibility for the direct contribution of Dnmt3a depletion in hematopoietic cells to development of PAH. The spontaneous occurrence of PAH and RVF in *Dnmt3a^-/-^* mice, along with their exacerbation under transient hypoxic conditions, provides robust evidence that *DNMT3A* mutations are sufficient to precipitate PAH and show that environmental factors may accelerate or exacerbate DNMT3A-associated PAH.

Epigenetic mechanisms, encompassing modifications in DNA methylation, histone acetylation, and the production of micro-RNAs, serve as a bridge between genetic and environmental factors^42^. Our team has demonstrated that excessive DNA methylation of specific target genes plays a role in the development of PAH^43–45^. Recent research indicates that altered function of TET2 and DNMT3A, enzymes regulating DNA demethylation and methylation respectively, may primarily regulate different genes^14^, perhaps explaining how both *TET2* mutations and *DNMT3A* mutations, though having opposing effects on methylation, have a concordant effect on inflammation. Our findings further demonstrate increased adverse remodeling of the pulmonary vasculature and reduced RV function in mice lacking Dnmt3a (**Figures 4-5**), which aligns with previous evidence implicating DNMT3A deficiency in the development of atherosclerotic lesions and cardiac dysfunction^46,47^. Notably, we confirmed that *Dnmt3a^-/-^* mice had normal LV function and no systemic hypertension. Depletion of Dnmt3a in hematopoietic cells led to heightened lung inflammation, notably the accumulation of macrophages (**Figure 6**)^48^. Macrophages are recognized contributors to vascular remodeling in PAH, likely becuase they can stimulate the proliferation of smooth muscle cells, leading to adverse pulmonary artery remodeling^48,49^. Interleukin-13 (IL-13) was also significantly higher in the plasma of *Dnmt3a^-/-^*mice. This cytokine is associated with macrophage/monocyte activation in humans and mice with PAH^43^. The mouse experiments provide direct evidence linking hematopoietic Dnmt3a depletion to adverse vascular remodeling and inflammation, two prominent characteristics of PAH^47^.

*TET2*-and *DNMT3A*-mutations cause a shift towards a pro-inflammatory macrophage phenotype^4445^. In a prior study using hematopoietic *Tet2* knockout mice (which developed PAH), we found that the IL-1ß antibody, Canakinumab, did regress PAH and reduce pulmonary vascular disease^20^. In this study, although there was no detectable difference between IL-1β levels in the plasma of control and *Dnmt3a^-/-^*mice, macrophages (the main producers and secretors of IL-1β) were the most predominant immune cells in the lung tissue of *Dnmt3a^-/-^* mice. Consistent with this, we observed adverse pulmonary vascular remodeling (and normal left heart function) and showed the lungs were inflamed, offering evidence that the inflammation caused the adverse pulmonary remodeling. Consistent with the pathologic role of inflammation in these mice, canakinumab regressed PAH in *Dnmt3a^-/-^* mice. Further work is required to assess the effect of canakinumab at the tissue level and on the lungs’ immune profile of these mice. Interestingly, there was no sex difference in susceptibility to PAH in mice.

Our study differs from the NIHR BioResource-Rare Diseases PAH study^46^, which did not identify this mutation as a human PAH gene, in several important aspects. Firstly, while their study focused on investigating deleterious genetic variations in 1048 patients with IPAH and familial PAH (FPAH), it did not include patients with APAH. In contrast, our study encompassed a larger sample size (∼ 2.5 times greater), consisting of 2572 patients, and specifically included APAH patients, who comprised 48% of all PAH patients in our dataset. Secondly, the methodology employed in our study differed as well. Instead of conducting whole-genome sequencing (WGS) on the PAH Biobank patients, as done in the NIHR BioResource-Rare Diseases study, we utilized whole-exome sequencing, which focuses on the protein-coding regions of the genome. Our approach allows for a more targeted analysis of potential disease-related genetic variations. Additionally, our study incorporated higher read depth, approximately 2-3 times greater than the previous study, which enhances the accuracy and reliability of variant detection. Further, we also investigated somatic mutations using targeted, high-depth, sequencing, a crucial difference in our study, since somatic mutations are more likely to drive CHIP-related disease development and progression and are commonly found at depths that may elude the sensitivity of WGS or WES.

### Study Limitations

The study has several limitations. First, panel sequencing of *DNMT3A* was only completed in a subset of the PAH Biobank cohort. Expanding the panel sequencing to include the entire PAH Biobank cohort will likely identify even more CHIP mutations, enhancing our statistical power to identify DNMT3A somatic mutations as a significant PAH CHIP gene. Second, since blood-related disorders are often associated with CHIP mutations, we screened for, and found, a small number of patients with hematologic conditions. We identified 6 *DNMT3A-*mutant patients with anemia and 1 with thrombocytopenia. This is consistent with prior studies showing that although CHIP mutations promote leukemia, the absolute risk is relatively low, only 0.5-1% per year^47^. Furthermore, CHIP has been found to increase risk of death by 40%^47^, though one study noted that only 1/246 CHIP carriers died from hematologic cancer^48^, suggesting cardiovascular disease may present a more serious conseqeunce of CHIP than leukemia. Third, six of the DNMT3A CHIP mutant patients identified by WES (n=2572) also underwent targeted panel sequencing (n=710). Panel sequencing discovered CHIP in 5/6 of these previously detected cases, suggesting that panel sequencing is adequately identifying the samples detected by WES, and more. We are performing deep panel sequencing on the remainder of the Biobank samples. Fourth, the experiments involving mice, we faced limitations in terms of the small sample size, which hindered our ability to optimally separate the data based on sex. As a result, after seeing no distinct sex differences, we made the decision to combine the female and male data together.

## Conclusion

This study expands our understanding of the genetic factors contributing to PAH. While *DNMT3A* mutations occur in IPAH they are most prevalent in female patients with APAH. To the best of our knowledge, this is the first gene mutation found to be associated with APAH. We observed distinct expression patterns of the three DNMT3A isoform transcripts, which indicates a potential mechanism of gene regulation through alternative splicing and/or epigenetic regulation. These findings contribute to a deeper understanding of the molecular mechanisms underlying PAH and underscore the potential utility of DNMT3A expression as a promising biomarker and therapeutic target for PAH.

## Source of funding

This study was supported in part by U.S. National Institutes of Health (NIH) grants NIH 1R01HL113003-01A1 (S.L.A.), NIH 2R01HL071115-06A1 (S.L.A), NIH R24HL105333 (W.C.N., M.W.P.), NIH R01HL160941 (W.C.N., M.W.P.), Canada Foundation for Innovation 229252 and 33012 (S.L.A.), Tier 1 Canada Research Chair in Mitochondrial Dynamics and Translational Medicine 950-229252 (S.L.A.), Canadian Institutes of Health Research (CIHR) Foundation Grant CIHR FDN 143261, the William J. Henderson Foundation (S.L.A.), and Canadian Vascular Network Scholar Award (L.T.; F.P.), and the JPB Foundation (W.K.C.). Further support was provided by a Vanier Canada Graduate Scholarship and CIHR MD/PhD Studentship (E.C.), an Ontario Molecular Pathology Research Network (OMPRN)/ Ontario Institute for Cancer Research (OICR) Cancer Pathology Translational Research Grant, CIHR Project Grant 451147, and a Canada Foundation for Innovation Grant (M.J.R.).

| **Nonstandard Abbreviations and Acronyms** |  |
| --- | --- |
| **APAH** | Associated Pulmonary Arterial Hypertension |
| **CHIP** | Clonal Hematopoiesis of Indeterminate Potential |
| **D-MIS** | Deleterious Missense Variants |
| **GEO** | Gene Expression Omnibus |
| **IPAH** | Idiopathic Pulmonary Arterial Hypertension |
| **DNMT3A** | DNA Methyltransferase 3A |
| **LGD** | Likely-Gene-Disrupting variants |
| **LHC** | Left Heart Catheterization |
| **MAPSE** | Mitral Annular Plane Systolic Excursion |
| **PAAT** | Pulmonary Artery Acceleration Time |
| **PAH** | Pulmonary Arterial Hypertension |
| **PCWP** | Pulmonary Capillary Wedge Pressure |
| **PVR** | Pulmonary Vascular Resistance |
| **RHC** | Right Heart Catheterization |
| **ROC** | Receiver Operating Characteristic |
| **RVF** | Right Ventricular Failure |
| **RVSP** | Right Ventricular Systolic Pressure |
| **TAPSE** | Tricuspid Annular Plane Systolic Excursion |
| **TET2** | Tet methylcytosine dioxygenase 2 |
| **WGS** | Whole Genome Sequence |

## Data Availability

All data are available in response to appropriate written requests.

## Acknowledgments

The authors would like to acknowledge the technical and scientific support of the Queen’s Cardiopulmonary Unit (QCPU), particularly Brooke Ring, Dr. Charles Hindmarch and Oliver Jones. Samples and/or Data from the National Biological Sample and Data Repository for PAH, which receives government support under an investigator-initiated grant (R24 HL105333) awarded by the National Heart Lung and Blood Institute (NHLBI), were used in this study. We thank contributors, including the Pulmonary Hypertension Centers who collected samples used in this study, as well as patients and their families, whose help and participation made this work possible. We thank and acknowledge the contribution of Russel Hirsch MD, Michelle Cash, S. Melissa Magness, Mukta Barve, Cincinnati Children’s Hospital Medical Center, Cincinnati, OH, United States; R. James White MD, PhD, Alison Light, Alison Theuer, University of Rochester Medical Center, Rochester NY, United States; Marc Simon MD, Traci McGaha, University of Pittsburgh, Pittsburgh PA, United States; David Badesch MD, Holly del Junco, Lisa Nicotera, Kelly Hannon, University of Colorado Denver, Aurora CO, United States; Erika Rosenzweig MD, Daniela Brady, Columbia University, New York NY, United States; Charles Burger MD, Inna Abrea, Andrea Tavlarides, Mayo Clinic Florida, Jacksonville FL, United States; Murali Chakinala MD, Sharon Heuerman, Washington University, St. Louis MO, United States; Thenappan Thenappan MD, Gretchen Peichel, Gina Paciotti, Brenda Vang, University of Minnesota, Minneapolis MN, United States; Greg Elliott MD, David Tomer, Quinn Montgomery, Department of Medicine at Intermountain Medical Center and the University of Utah, Murray UT, United States; Hap Farber, MD, Robert Simms MD, Eric Stratton, Boston University School of Medicine, Boston MA, United States; Robert Frantz MD, Louise Durst, Kristal Rohwer, Mayo Clinic, Rochester MN, United States; Jean Elwing MD, Tammy Roads, Autumn Studer, University of Cincinnati, Cincinnati OH, United States; Nicholas Hill MD, Karen Visnaw, Tufts Medical Center, Boston MA, United States; Dunbar Ivy MD, Kathleen Miller-Reed, Karlise Lewis, Children’s Hospital of Colorado, University of Colorado Denver, Aurora CO, United States; James Klinger MD, Amy Palmisciano, Meghan Ahearn, Rhode Island Hospital, Providence RI, United States; Steven Nathan MD, Merte Lemma, Inova Heart and Vascular Institute, Falls Church VA, United States; Ronald Oudiz MD, Joy Beckmann, Bindu John, LA Biomedical Research Institute at Harbor-UCLA, Torrance CA, United States; Ivan Robbins MD, Shannon Cordell, Vanderbilt University Medical Center, Nashville TN, United States; Robert Schilz DO, PhD, Mary Andrews, University Hospital of Cleveland, Cleveland OH, United States; Terry Fortin MD, Karla Kennedy, Susana Almeida-Peters, Duke University Medical Center, Durham NC, United States; Jeffrey Wilt MD, Kimberly McClain, Spectrum Health Hospitals, Grand Rapids MI, United States; Delphine Yung MD, Anne Davis, Linnea Brody, Seattle Children’s Hospital, Seattle WA, United States; Eric Austin MD, Karen Chaffin, Vanderbilt University-Peds, Nashville TN, United States; Ferhaan Ahmad MD, PhD, Page Scovel, Division of Cardiovascular Medicine, University of Iowa, Iowa City IA, United States; Nitin Bhatt MD, Joseph Santiago, Ohio State University, Columbus OH, United States. We would like to thank Cassianne Robinson-Cohen PhD, Department of Medicine at Vanderbilt University Medical Center, McGill University, and Calvin Sjaarda PhD for providing the human control sample for deep panel sequencing, S. Jaiswal, MD, University of Stanford, United States for kindly providing the flox mice for the study.

