## Supplementary material for "Germline and Somatic Mutations in DNA Methyltransferase 3A *(DNMT3A)* Predispose to Pulmonary Arterial Hypertension (PAH) in Humans and Mice: *Implications for Associated PAH*": Al-Qazazi, Emon et al. 2023 Supplemental Data.docx

| **Table S1. Rare, predicted deleterious likely germline variants in candidate PAH risk gene *DNMT3A* among 2572 PAH Biobank cases.** | | | | | | | | | | | | | | |
| --- | --- | --- | --- | --- | --- | --- | --- | --- | --- | --- | --- | --- | --- | --- |
| **Patient ID** | **Nucleotide change** | **Amino acid change** | **MAF, gnomAD WES** | **CADD phred** | **REVEL score** | **PAH class** | **Genetic ancestry** | **Sex** | **Mean PAP, at rest (mmHg)** | **Mean PCWP, at rest (mmHg)** | **Cardiac output, Fick (L/min)** | **PVR, Fick (Wood units)** | **Mean SAP (mmHg)** | **Mean SAP:PAP** |
| 1. **04-046** | c.1471G>T | p.Glu491✶ | . | 38 | . | IPAH | Hispanic | F | 36 | 8 | 4.6 | 6.09 | NA | NA |
| 2. **29-005** | c.2083-1G>C | splicing | . | 25 | . | IPAH | EUR | F | 25 | 7 | NA | NA | 97 | 3.9 |
| 3. **07-060** | c.875T>C | p.Ile292Thr | 4.08E-06 | 24 | 0.72 | FPAH | EUR | F | 65 | NA | 3.4 | NA | 93 | 1.4 |
| 4. **12-147** | c.2711C>T | p.Pro904Leu | 3.25E-05 | 29 | 0.94 | FPAH | EUR | F | 69 | 13 | 4 | 14 | 75 | 1.1 |
| 5. **21-044** | c.2204A>G | p.Tyr735Cys | 4.51E-05 | 27 | 0.9 | APAH-Porto | EUR | F | 55 | 15 | 6.7 | 5.97 | NA | NA |
| 6. **13-025^†^** | c.1937-2A>G | splicing | . | 24 | . | APAH-Porto | EUR | M | 36 | 13 | NA | NA | NA | NA |
| 7. **22-056** | c.1903C>T | p.Arg635Trp | 5.99E-05 | 35 | 0.84 | APAH-HIV | EUR | F | 48 | 4 | 3.3 | 13.33 | NA | NA |
| 8. **03-053** | c.893G>A | p.Gly298Glu | 4.07E-06 | 30 | 0.87 | APAH-CTD | EUR | F | 43 | 5 | 2.2 | 17.27 | 94 | 2.2 |
| 9. **07-052** | c.2032C>T | p.Gln678✶ | . | 40 | . | APAH-CTD | EUR | F | 26 | 14 | 3.9 | 3.08 | 115 | 4.4 |
| 10. **20-010** | c.2204A>G | p.Tyr735Cys | 4.51E-05 | 26 | 0.9 | APAH-CTD | Hispanic | F | 55 | 14 | 4.1 | 10 | NA | NA |
| 11. **07-024** | c.2655G>C | p.Arg885Ser | . | 27 | 0.9 | APAH-CTD | EUR | F | 29 | 5 | NA | NA | NA | NA |
| 12. **06-101** | c.656A>G | p.Lys219Arg | . | 25 | 0.53 | APAH-CHD | African | F | 37 | 5 | 4.3 | 7.44 | 87 | 2.3 |
| ***Mean ± SD, DNMT3A carriers***  ***n, DNMT3A carriers***  ***Mean ± SD, cohort APAH+IPAH excluding DNMT3A carriers***  ***n, cohort APAH+IPAH excluding DNMT3A and TET2 carriers***  ***p-Value*** | | | | | | | | 11:1 | 44 ± 15 | 9 ± 4 | 4.1 ± 1.2 | 9.6 ± 4.9 | 94 ± 13 | 2.6 ± 1.3 |
|  |  |  |  |  |  |  |  |  | 12 | 11 | 9 | 8 | 6 | 6 |
|  |  |  |  |  |  |  |  |  | 49 ± 7 | 12 ± 4 | 6.1 ± 3.4 | 11 ± 8.1 | 96 ± 20 | 2.0 ± 0.7 |
|  |  |  |  |  |  |  |  |  | 2331 | 2270 | 1680 | 1623 | 1470 | 1412 |
|  |  |  |  |  |  |  |  |  | NS | NS | NS | NS | NS | NS |

✶*DNMT3A* transcript: NM_175629.2;. **_†_** Patients carry more than one variant in the candidate risk genes. **Abbreviations:** **APAH-CHD**, pulmonary arterial hypertension associated with congenital heart disease; **APAH-CTD**, pulmonary arterial hypertension associated with connective tissue diseases; **APAH-**Porto, pulmonary arterial hypertension associated with porto-pulmonary hypertension; **FPAH**, familial pulmonary arterial hypertension; **MAF**, minor allele frequency; **IPAH**, idiopathic pulmonary arterial hypertension; **PAP**, pulmonary artery pressure; **PCWP**, pulmonary capillary wedge pressure; **PVR**, pulmonary vascular resistance; **SAP**, systemic arterial pressure**; WES**, whole exome sequencing.

| **Table S2. Rare, predicted deleterious likely somatic variants in candidate PAH risk gene, *DNMT3A*, among 2572 PAH Biobank cases.** | | | | | | | | | | | | | | | | |
| --- | --- | --- | --- | --- | --- | --- | --- | --- | --- | --- | --- | --- | --- | --- | --- | --- |
| **Patient ID** | **Nucleotide change** | **Amino acid change** | **MAF, gnomAD WES** | **Alternate Allele Fraction** | **CADD phred** | **REVEL score** | | **PAH class** | **Sex** | **Genetic ancestry** | **Mean PAP, at rest (mmHg)** | **Mean PCWP, at rest (mmHg)** | **Cardiac output, Fick (L/min)** | **PVR, Fick (Wood units)** | **Mean SAP (mmHg)** | **Mean SAP:PAP** |
| **03-068** | c.1226G>T | p.Trp409Leu | . | 22% | 34 | 0.75 | | IPAH | F | EUR | 50 | 7 | 3.8 | 11.44 | 97 | 1.94 |
| **22-027^†^** | c.2644C>T | p.Arg882Cys | 1.00E-04 | 42% | 34 | 0.89 | | IPAH | F | EUR | 50 | 6 | 3.7 | 11.89 | NA | NA |
| **15-051** | c.1258A>T | p.Lys420* | . | 14% | 38 | . | | IPAH | F | EUR | 95 | 9 | NA | NA | 61 | 0.64 |
| **03-061^††^** | C.976C>T | p.Arg326Cys | 4.47E-05 | 18% | 33 | 0.77 | | IPAH | M | Hispanic | 57 | 12 | 6.9 | 6.52 | 107 | 1.9 |
| **05-202** | c.2174-2A>G | splicing | 9.37E-06 | 23% | 24 | . | | IPAH | F | EUR | 35 | 14 | 6.8 | 3.09 | 102 | 2.9 |
| **12-102** | c.2200T>C | p.Phe734Leu | . | 13% | 32 | 0.90 | | FPAH | M | EUR | 42 | 4 | NA | NA | NA | NA |
| **13-067** | c.2026C>T | p.Arg676Trp | 4.49E-05 | 13% | 35 | 0.91 | | DTOX | F | African | 45 | 18 | 6.8 | 3.97 | NA | NA |
| **12-041** | c.2116G>A | p.Gly706Arg | . | 19% | 33 | 0.97 | | DTOX | M | EUR | 50 | 12 | NA | NA | 97 | 1.94 |
| **05-168^†^** | c.2645G>A | p.Arg882His | 2.00E-04 | 28% | 33 | 0.74 | | APAH-Porto | M | EUR | 66 | 9 | 5.1 | 11.18 | 98 | 1.5 |
| **13-021^†^** | c.2645G>A | p.Arg882His | 2.00E-04 | 28% | 33 | 0.74 | | APAH-CTD | F | EUR | 35 | 11 | 6.9 | 3.48 | NA | NA |
| **16-033** | c.920C>G | p.Pro307Arg | 8.96E-06 | 13% | 27 | 0.97 | | APAH-CTD | F | EUR | 32 | 10 | NA | NA | 72 | 2.25 |
| **22-067^††^** | c.1742_1743delinsAT | p.Trp581Tyr | 4.09E-06 | 18% | 43 | . | | APAH-CTD | F | Hispanic | 35 | 13 | NA | NA | NA | NA |
| **22-013** | c.2206C>T | p.Arg736Cys | 3.28E-05 | 20% | 34 | 0.92 | | APAH-CTD | F | EUR | 32 | 8 | 7.1 | 3.38 | 67 | 2.09 |
| **12-180** | c.2221G>C | p.Arg741Pro | . | 23% | 25 | 0.83 | | APAH-CTD | F | EUR | 60 | 12 | NA | NA | NA | NA |
| **12-192** | c.2402T>C | p.Met801Thr | . | 12% | 27 | 0.93 | | APAH-CTD | F | EUR | 55 | 6 | 4.0 | 12.31 | 99 | 1.8 |
| **11-057^††^** | c.2645G>A | p.Arg882His | 0.0002 | 19% | 33 | 0.74 | | APAH-CTD | F | EUR | 60 | 11 | 4.9 | 10 | 122 | 2 |
| **28-039** | c.2695C>T | p.Arg899Cys | 6.54E-05 | 24% | 34 | 0.94 | | APAH-CTD | F | EUR | 54 | 7 | NA | NA | NA | NA |
| **24-008** | c.1258A>T | p.Lys420* | . | 14% | 38 | . | | APAH-CHD | F | EUR | 49 | 9 | 1.8 | 21.98 | 47 | 0.96 |
| **29-037^†^** | c.2644C>T | p.Arg882Cys | 1.00E-04 | 39% | 34 | 0.89 | | APAH-CHD | F | EUR | 88 | 16 | 3.9 | 18.46 | 93 | 1.1 |
| **08-111^††^** | c.1903C>T | p.Arg635Trp | 5.99E-05 | 10% | 35 | 0.84 | | APAH | F | Hispanic | NA | NA | NA | NA | NA | NA |
| **17-004^††^** | c.1937-2A>G | splicing | . | 15% | 24 | . | | APAH | F | EUR | NA | NA | NA | NA | NA | NA |
| **Mean ± SD, DNMT3A carriers** | | | |  |  |  |  | | 4.25:1 |  | 52 ± 17 | 10 ± 4 | 5.1 ± 1.7 | 9.8 ± 6.1 | 89 ± 22 | 1.8 ± 0.6 |
| **n, DNMT3A carriers** | | | |  |  |  |  | |  |  | 19 | 19 | 12 | 12 | 12 | 12 |

**DNMT3A* transcript: NM_175629.2. **^†^**Exceptions to the mosaic pipeline variant filter (MAF and alternate allele fraction); detected as known mutation hotspot in cancer, ††Exceptions to the mosaic pipeline variant filter (MAF or minimum read depth); detection by the germline pipeline with posterior odds >10. Variant filter: allele frequency <0.0001 and likely gene disrupting (stop/gain, frameshift, or canonical splicing) or missense with REVEL score >0.5. **Abbreviations:** APAH-CHD, pulmonary arterial hypertension associated with congenital heart disease; APAH-CTD, pulmonary arterial hypertension associated with connective tissue diseases; APAH-Porto, pulmonary arterial hypertension associated with portopulmonary hypertension; FPAH, familial pulmonary arterial hypertension; MAF, minor allele frequency; IPAH, idiopathic pulmonary arterial hypertension; PAP, pulmonary artery pressure; PCWP, pulmonary capillary wedge pressure; PVR, pulmonary vascular resistance; SAP, systemic arterial pressure; WES, whole exome sequencing; PAP, pulmonary artery pressure; PCWP, pulmonary capillary wedge pressure; PVR, pulmonary vascular resistance; SAP, systemic arterial pressure

| **Table S3: Response to Vasodilators* and other PAH medication among DNMT3A deleterious variant carriers** | | | |
| --- | --- | --- | --- |
| **Vasodilators** | **APAH+IPAH excluding DNMT3A and TET2 carriers** | | **APAH+IPAH DNMT3A carriers** |
| **Oxygen + Nitric Oxide** | 50/283 (17.7%) | | 0/3 (0%) |
| **IV Epoprostenol** | 13/105 (12.4%) | | 0/5 (0%) |
| **Inhaled Nitric Oxide** | 60/559 (10.7%) | | 0/5 (0%) |
| **Oxygen** | 5/16 (31.2%) | | - |
| **IV Nitroprusside** | 1/7 (14.3%) | | - |
| **IV Adenosine** | 5/38 (13.1%) | | - |
| **Inhaled Iloprost** | 1/2 (50%) | | - |
| **Inhaled Epoprostenol** | 1/6 (16.7%) | | - |
| **Alprostadil** | 2/8 (25%) | | - |
| **Calcium Channel Blocker** | 0/3 (0%) | | - |
| **Unknown** | 2/16 (12.5%) | | - |
| **Total** | 140/1043 (13.4%) | | 0/13 (0%) |
| **PAH Medications** | **APAH+IPAH excluding DNMT3A and TET2 carriers** | | **APAH+IPAH DNMT3A carriers** |
| **PDE5 inhibitor** | | 2004/2664 (75.2%) | 10/10 (100%) |
| **Prostacyclin analog** | | 1352/2664 (50.7%) | 4/10 (40%) |
| **Endothelin receptor inhibitor** | | 1031/2664 (38.7%) | 5/10 (50%) |
| **Stimulator of soluble guanylate cyclase** | | 43/2664 (1.6%) | 1/10 (10%) |
| **Calcium channel blocker** | | 239/2664 (9 %) | 0/10 (0%) |
| **RTK inhibitor** | | 5/2664 (0.2%) | 0/10 (0%) |
| **Unknown** | | 29/2664 (1.1%) | 0/10 (0%) |

* Positive vasodilator responders were defined according to standard criteria as a drop of mean pulmonary arterial pressure (mPAP) >10mmHg to mPAP at rest <40mmHg with preserved or improved cardiac output. **Abberviations:** APAH, associated PAH; IPAH, idiopathic PAH; IV, intra-venous; PDE5, phosphodiesterase 5; RTK, receptor tyrosine kinase

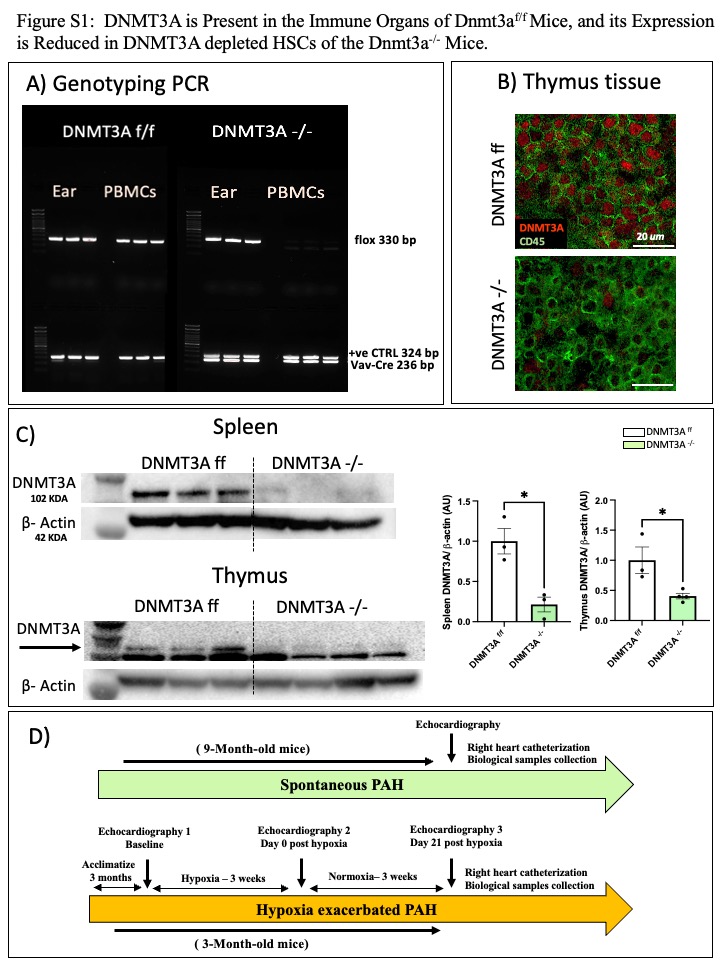

**Figure S2:** Showing no significant change between controls (*Dnmt3a^f/f^*) and *Dnmt3a^-/-^ mice* in LVSP, LVEDP and MAPSE measured using open chest LV catheterization (LVSEP, LVEDP) and cardiac ultrasound (MAPSE).

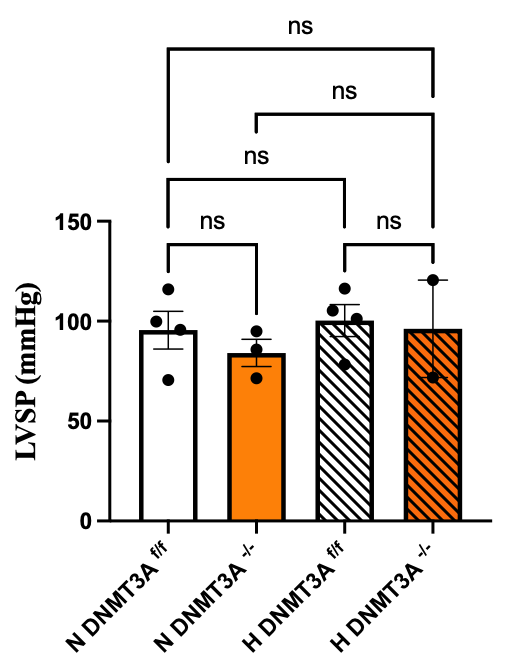

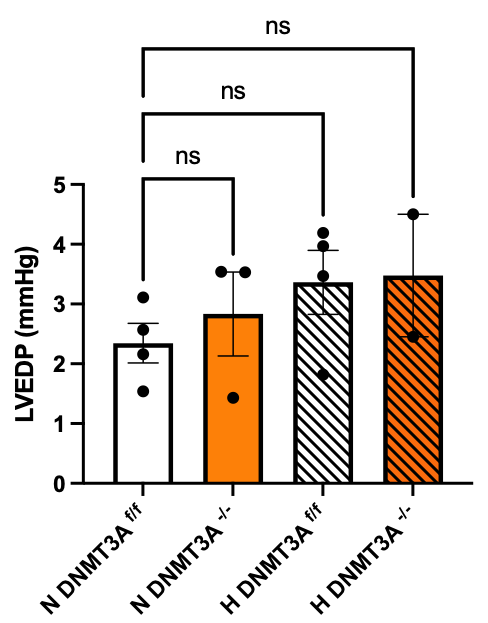

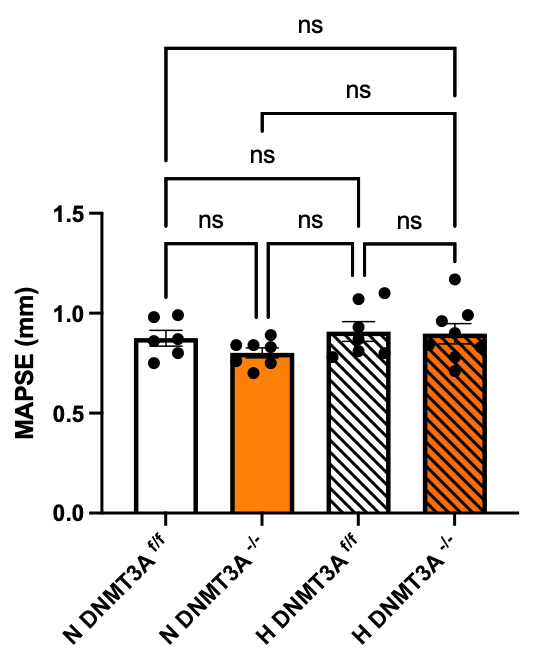

**Figure S3:** Showing differences between percentage of CD45+ cells that are neutrophils, T-cells and B-cells in the lungs of *Dnmt3a^f/f^* and *Dnmt3a^-/-^* mice obtained using flow cytometry.

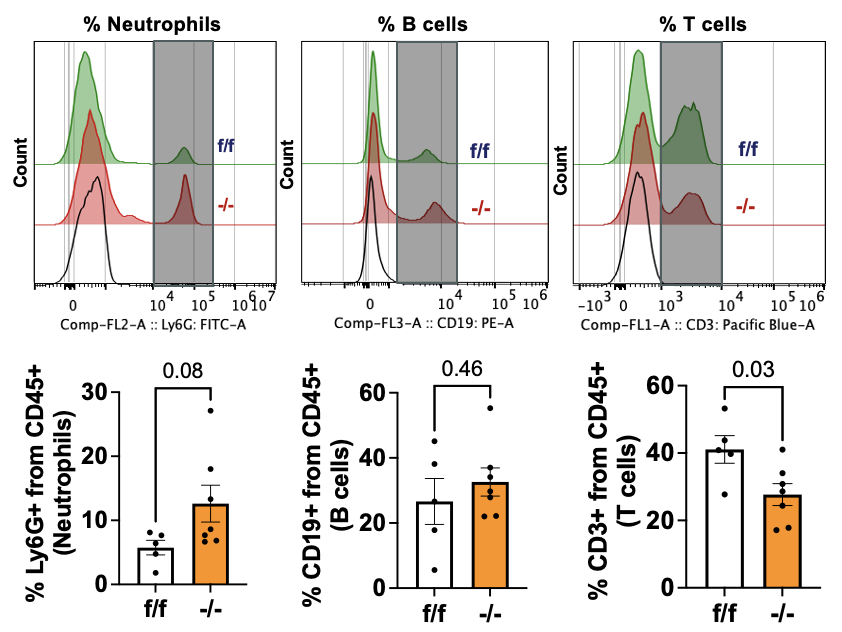
