## Supplementary material for "Germline and Somatic Mutations in DNA Methyltransferase 3A *(DNMT3A)* Predispose to Pulmonary Arterial Hypertension (PAH) in Humans and Mice: *Implications for Associated PAH*": Al-Qazazi, Emon et al. 2023 Supplemental Methods.docx

1- **Immunohistochemistry**

Paraffin sections (5 µm) were deparaffinized and hydrated. Antigen retrieval was achieved by microwave heating using a citrate buffer (10 mM, pH 6.0; 20 min). Following antigen retrieval, sections were allowed to cool and washed with PBS-tween 0.05% (PBS-T). For immunofluorescence staining, sections were blocked with 1 % Bovine Serum Albumin (BSA) for 30 min at room temperature. Sections were incubated overnight with primary antibodies (CD45) at 4^o^C (**Supplemental Table 4**). Next, sections were washed with PBS-T and incubated with conjugated secondary antibodies (**Supplemental** **Table** **4**) for 1-hour at room temperature. Finally, slides were washed and mounted using ProLong Gold Antifade Mounting with DAPI (ThermoFisher; Waltham, MA, USA). The fluorescence-based imaging was performed using TCS SP8 laser scanning confocal microscope (SP8 Leica, Concord, ON, Canada) and the HC PL APO CS2 63x/1.40 oil objective.

2- **Hematoxylin and Eosin (H&E)**

Paraffin sections of RV tissue and lung were deparaffinized and hydrated. Sections were stained using hematoxylin and eosin (H&E Staining Kit ab245880, Abcam). Morphometric analysis was performed to measure pulmonary arterial wall thickness. The artery medial wall thickness was estimated by measuring the outer and inner layers of arteries in the transversal section, and it was calculated as follow: % Artery medial wall thickness = [1-(perimeter of inner layer/perimeter of outer layer)]*100%.

3- **Collagen deposition quantification**

Paraffin sections of RV and LV tissue were deparaffinized and hydrated. The nuclei were stained with Weigert’s hematoxylin and subsequently stained with Picrosirius Red Stain as per the manufacturer's instruction (Sigma Aldrich; “Direct Red 80” Cat#365548).

Images were acquired using Leica DM 4000 microscope equipped with a DFC310 FX camera using a 20x objective and LASX software. The collagen fibers (defined by their red stain) were quantified using ImageJ by measuring the percentage of the red signal in the field.

4- **Western Blotting**

Total Thymus and spleen protein was prepared in anti-protease buffer (#9803; Cell Signaling Technologies, Beverly, MA, United States) and measured using the BCA protein reagent assay (A/B) (Thermo scientific; cat# 23228, 1859078). 50ug (thymus) and 80 ug (spleen) of protein were loaded on a 4-12% gradient polyacrylamide gel for electrophoretic separation using the SDS-PAGE method. Running buffer was prepared using 20X MOPS Buffer (novex: REF NP0001), 0.1% Bolt Antioxidant (Invitrogen: REF BT0005) and ddH_2_O. Resolved proteins were transferred to polyvinylidene fluoride (PVDF) membranes (Novex cat# LC2002) and blocked in 5% milk. Immunoblotting was performed using the primary antibodies as described in **Supplemental** **Table** **4**. Membranes were developed using ECL Western blot detection reagent (Amersham: REF RPN2232), imaged using ChemiDoc^TM^MP imaging system (Bio-Rad Laboratories; Mississauga, ON, Canada) and analyzed with ImageJ.

5- **Flow cytometry**

Lung single-cell suspensions were obtained using organ-specific protocols. Upon animal euthanasia, the lungs were harvested and kept in a calcium-free Hanks medium (Sigma Aldrich) until processing. The lungs were minced and digested using DNase (100 mg/mL) and Liberase (13 WU/mL) for 30 minutes at 37^o^C (2 cycles of 15 minutes). The cell suspensions of lung were filtered through a 70 µm cell strainer, and the total cell number was calculated (10^5^ cells per tube). The LIVE/DEAD™ Fixable Near-IR Dead Cell Stain Kit (ThermoFisher Scientific L10119) was used to discriminate dead cells, as per manufacturers’ recommendation. Cells were stained using fluorescent-tagged antibodies against surface antigens (**Supplemental Table 4**) and washed with FACS buffer. Cells were fixed with 2% PFA (15 minutes, 4^o^C). The SH-800S cell sorter/cytometer (Sony Biotechnology San Diego, CA, USA) was used to acquire the cytometric data. The data was analyzed using the FlowJo^TM^ (BD Life Sciences, USA).

**Supplemental Table 4: Antibodies used for mice experiments.**

| Antibody | Technique | Working dilution | Cat#/Manufacturer |
| --- | --- | --- | --- |
| Primary antibodies | | | |
| Rabbit DNMT3A | Western blot  Immunofluorescence | 1:1000  1:50 | Abcam- AB307503 |
| Rat anti CD45 | Immunofluorescence | 1:100 | Invitrogen – 14-0451-82 |
| Leukocytes CD45 | Flow Cytometry | 1:100 | Biolegend -103127 |
| MacrophagesF4/ 80 | Flow Cytometry | 1:100 | Biolegend - 157305 |
| B cells CD19 | Flow Cytometry | 1:100 | Biolegend - 152407 |
| Neutrophils LY6G | Flow Cytometry | 1:100 | Biolegend - 127605 |
| T cells CD3 | Flow Cytometry | 1:100 | Biolegend - 100213 |
| Secondary antibodies | | | |
| Alexa Fluor 647 goat-anti-rabbit IgG | Immunofluorescence | 1:400 | **Invitrogen -** A-21244 |
| Cy3 goat anti-rat | Immunofluorescence | 1:400 | Jackson Immuno Research Laboratories 112-165-003 |
| eBioscience™ 10X RBC Lysis Buffer | * | * | Thermo-Fisher 00-4300-54 |
| Liberase (13WU/ml) | * | * | Sigma Millipore (Roche) |
| DNase (100mg/ml) | * | * | Sigma Millipore -10104159001 |
